# The impact of population-wide rapid antigen testing on SARS-CoV-2 prevalence in Slovakia

**DOI:** 10.1101/2020.12.02.20240648

**Authors:** Martin Pavelka, Kevin Van-Zandvoort, Sam Abbott, Katharine Sherratt, Marek Majdan, CMMID COVID-19 working group, Inštitút Zdravotných Analýz, Pavol Jarčuška, Marek Krajčí, Stefan Flasche, Sebastian Funk

## Abstract

Slovakia conducted multiple rounds of population-wide rapid antigen testing for SARS-CoV-2 in late 2020, combined with a period of additional contact restrictions. Observed prevalence decreased by 58% (95% CI: 57-58%) within one week in the 45 counties that were subject to two rounds of mass testing, an estimate that remained robust when adjusting for multiple potential confounders. Adjusting for epidemic growth of 4.4% (1.1-6.9%) per day preceding the mass testing campaign, the estimated decrease in prevalence compared to a scenario of unmitigated growth was 70% (67-73%). Modelling suggests that this decrease cannot be explained solely by infection control measures, but requires the additional impact of isolation as well as quarantine of household members of those testing positive.

## Main Text

Non-pharmaceutical interventions have been extensively used worldwide to limit the transmission of SARS-CoV-2 (*1*). These have included travel restrictions, mandating face masks, closure of schools and non-essential businesses, and nationwide stay-at-home orders. While all the measures were aimed at mitigating ill-health due to COVID-19 (*2, 3*) they also place an unprecedented economic and social burden on people (*4, 5*), the majority uninfected. Testing of reported symptomatic cases and tracing their contacts aims to provide a more targeted measure but in many settings has proven insufficient for containing transmission (*6*).

Mass testing campaigns are an alternative way to identify infectious individuals and allow targeting of interventions without much added burden to those not infectious. However, they have been limited until recently by the dependence on Polymerase Chain Reaction (PCR) for the diagnosis of a SARS-CoV-2 infection. While laboratory capacities have been upscaled in record time, PCR testing remains expensive and often has turnaround times of more than one day, diminishing its utility (*7*). The PCR detection window also typically extends to the post-infectious period by detecting RNA fragments, hence identifying infected who are no longer infectious (*8*).

In comparison, recently developed rapid antigen tests are cheap and can be quickly produced in large quantities offering results on site in 15-30 mins without the need for a laboratory. They are less sensitive in detecting infections with low viral load but have been found to detect over 70% of likely infectious infections, making mass testing a viable part of the portfolio of non-pharmaceutical interventions (*9, 10*). A recent observational study estimated the sensitivity of lateral flow devices in detecting infectious individuals to be as high as 83-91% (*11*).

In October and November 2020, Slovakia used rapid antigen tests in a campaign targeting the whole population in order to identify infectious infections at scale, rapidly reduce transmission and allow quicker easing of lockdown measures (*12*). A pilot took place between 23 and 25 October in the four most affected counties, followed by a round of national mass testing on 31 October and 1 November (henceforth: round 1). High prevalence counties were again targeted with a subsequent round on 7 and 8 November (round 2).

In total, 5,276,832 SD-Biosensor Standard Q rapid antigen tests were conducted by trained medical personnel during the mass testing campaigns, with 65% of the respective populations tested in the pilot, 66% in mass testing round 1 and 62% round 2. This corresponded to 87%, 83% and 84% of the age-eligible population (10 and 65 years and older adults in employment) in each round, respectively, and does not include residents who were quarantining at the time of the campaign and another 534,300 tests that were conducted through additional testing sites dedicated to medical, military and governmental personnel and not included in geographical county data.

A total of 50,466 participants tested positive, indicating the presence of a currently infectious SARS-CoV-2 infection. The proportion of positive tests was 3.91% (range across counties: 3.12 to 4.84%) in the pilot, 1.01% (range: 0.13-3.22%) in round 1 and 0.62% (range: 0.28-1.65%) in round 2 (Figure 2C and D).

Potentially large numbers of false positive tests have been a point of criticism for mass testing campaigns. While multiple studies have found high specificity of the Biosensor test kit they are not powered to exclude specificity levels that on population level would yield an overwhelming amount of false positives (*13*). From the low test-positive rates in some counties, we estimate that with 95% certainty the specificity of the SD Biosensor Standard Q antigen test was exceeding 99.85% and therefore not of major concern in this study.

The counties with the highest prevalence were found in the Northern part of the country, while the two main Slovakian cities of Bratislava and Košice had some of the lowest observed prevalence (Figure 1c). Reflecting this pattern, we found high county-level prevalence to be associated with younger average population age and lower population density (Figure S8). Given that prevalence varied at a much smaller geographical length scale (*14*), such associations may be clearer at the individual or community level as observed in other countries.

**Figure 1:**
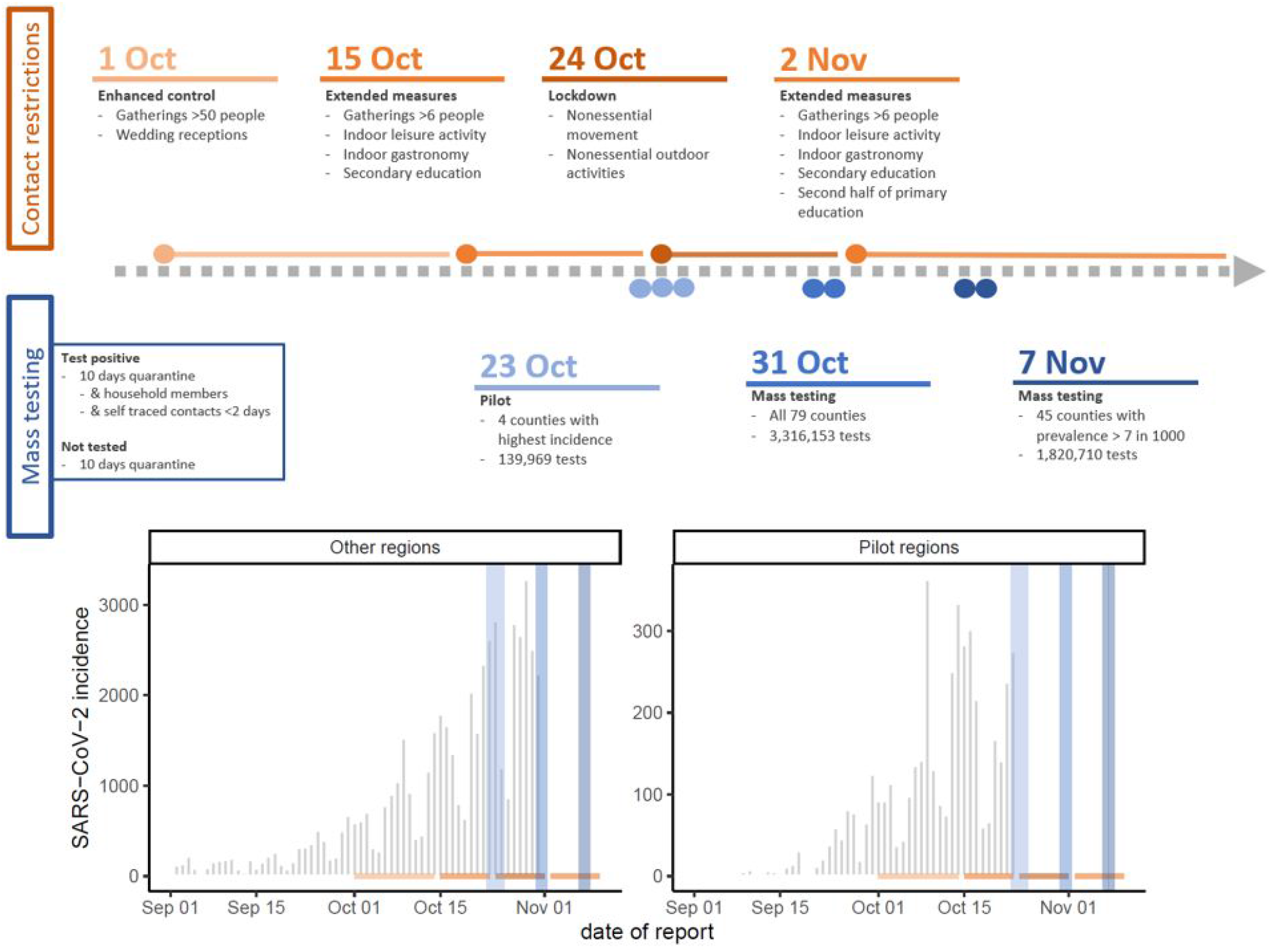
Overview of interventions and pre mass testing epidemiology. Top panel: description of timing and extent of national contact restriction in Slovakia (color intensity indicates intensity of the measures) and timing and extent of the mass testing campaigns. Dots and lines in respective colors show the start and duration of the contact restrictions and the blue dots show the days on which mass testing was conducted, though the highest turnout was usually on the first day. The additional box illustrates contact reducing measures for test positives and those who did chose not to get tested. Bottom panel: SARS-CoV-2 infection incidence as reported by the Slovak Ministry of Health and collected through passive symptom triggered PCR testing. Using the same color coding as in the top panel contact interventions are displayed by horizontal and mass testing campaigns by vertical lines. Data following the respective first mass testing campaign is omitted as mass testing is likely to have interfered with passive surveillance.

In the four counties where the pilot was conducted, observed infection prevalence decreased by 56% (95% Confidence Interval, CI: 54-58%) between the pilot and round 1 of the mass testing campaign and a further 60% (95% CI: 56-63%) between rounds 1 and 2, totalling a decrease of 82% (95% CI: 81-83%) over two weeks. There was little heterogeneity between counties (Figure 2B).

**Figure 2:**
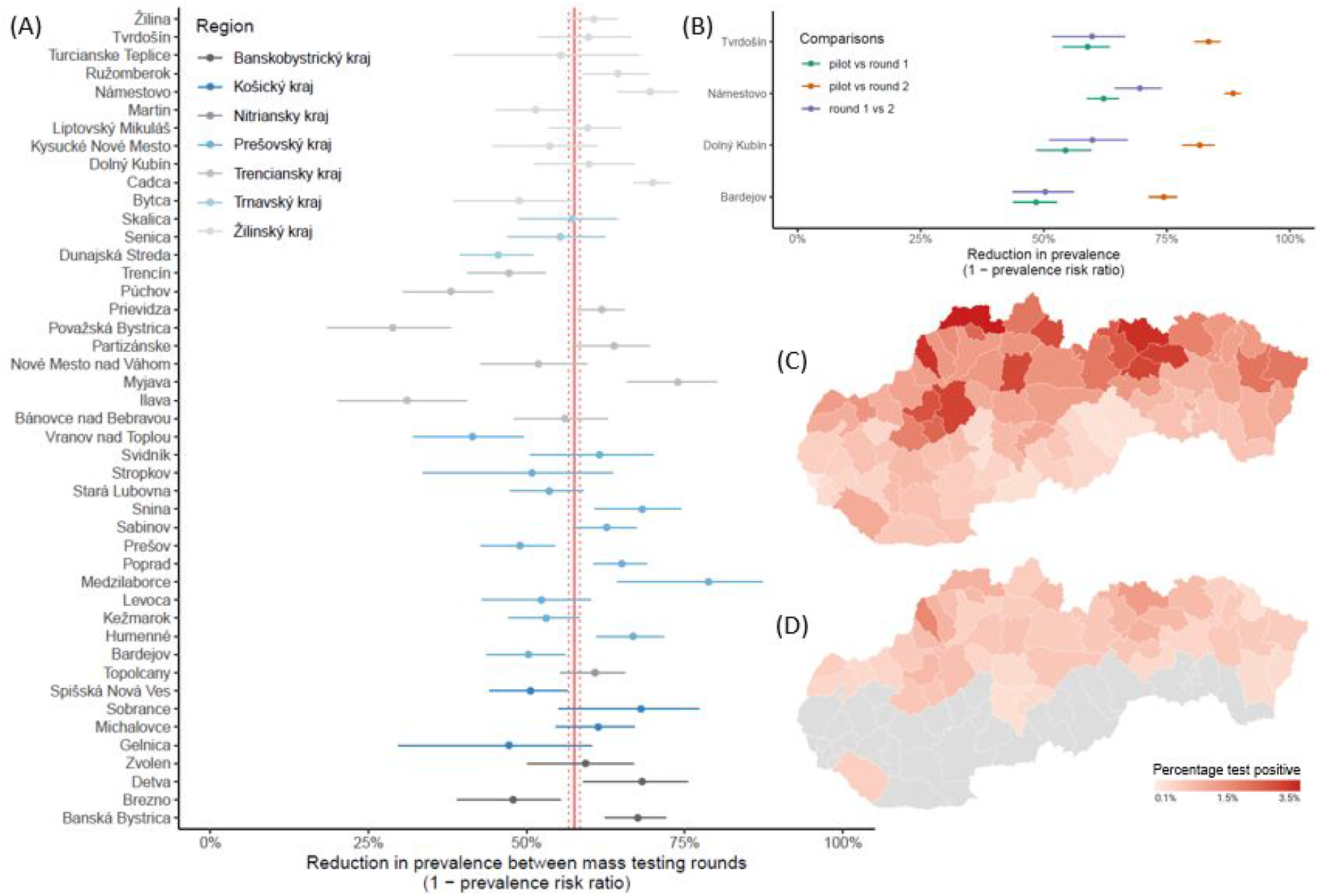
The change in test positivity between mass testing campaigns. Panel A: change in test positivity (1 - cPR) observed from mass testing round 1 to round 2 in the 45 counties that were eligible for both rounds of mass testing. Counties are grouped and color coded into regions. The crude pooled estimate and its 95% confidence bounds are shown as red vertical lines. Panel B: change in test positivity (1 - cPR) observed from the pilot mass testing round to either the first (green) or the second (orange) national round and from the first to the second mass testing round (blue) in the 4 counties that were included in the pilot. Panel C and D: county level test positivity in the first (C) and second (D) round of mass testing. Grey areas indicate counties that were not part of the second round because their test positivity rate was less than 7 per 1000 and hence have no estimates.

Among the 45 counties that were included in round 2 of the mass testing campaign, observed infection prevalence decreased by 58% (95% CI: 57-58%). Combining the pilot results with the ones from the two rounds of testing in 45 counties each round of mass testing was estimated to have reduced observed infection prevalence by 56% (95% CI: 52-59%) when adjusted for attendance rates, reproduction number and prevalence in previous rounds. The estimated reduction between rounds varied by county from 29% in county Považská Bystrica to 79% in county Medzilaborce but with little regional differences (Figure 2A). Neither region, attendance rates, prevalence in round 1 or the estimated growth rate prior to mass testing were found to be significantly associated with county specific reductions.

At the time of round 1 of the mass testing campaign, incidence of confirmed cases reported through the syndromic surveillance system was rising in non-pilot counties with an estimated infection growth rate of 4.4% (1.1%-6.9%) per day. When adjusting for this growth trend, we estimated a self-adjusted prevalence ratio (saPR) of 0.30 (0.27-0.33). In the pilot counties, reported infection incidence showed signs of levelling in the week before the mass testing campaign with an estimated infection growth rate of 1.3% (-7.4-7.8%), yielding a respective saPR of 0.31 (0.26-0.33).

As we used the test positivity rate of the subsequent round to estimate the impact of the previous one, we were unable to observe the full effect of the campaign. However, we find that the reduction achieved per round of testing was 56% (52-59%), indicating that the 41 counties with two rounds of testing likely reduced infection prevalence by 81% (77-83%) within two weeks and that the 4 counties included into the pilot testing reduced infection prevalence by 91% (89-93%) within three weeks.

The observational nature of this study made it difficult to clearly distinguish the effect of the mass testing campaigns from that of the other non-pharmaceutical interventions introduced at a similar time, that have led to a reduction in contacts and mobility, albeit much less than during the Spring lockdown (Figure S4). A reduction in greater than 50% decline in infection prevalence within one week (or 80% in two weeks) is striking, particularly while primary schools and workplaces were mostly open. For comparison, a month long lockdown in November in the UK resulted in just a 30% decrease in prevalence (*15*). This, alongside the inability to control the rebounding spread of SARS-CoV-2 in Slovakia through even more stringent contact restrictions in December, would suggest that a large share of the impact can be attributed to the mass testing campaigns.

In order to further investigate the relationship between the reduction in prevalence, mass testing and non-pharmaceutical interventions, we used a microsimulation model to simulate fine scale SARS-CoV-2 transmission in a representative county included in the pilot phase of the mass testing. Among the multiple intervention scenarios tested, only the scenario that assumed a substantial impact of both the additional contact reducing measures and the mass testing campaigns was able to generate reductions in test positivity rates between testing rounds that were similar to those observed (Figure 3). The requirement for quarantine for the whole household following a positive test was essential for the combined effect of mass testing and contact reduction measures; predicted prevalence ratio between the first two testing rounds of 0.30 (0.26-0.34) with and 0.78 (0.72-0.84) without household quarantine.

**Figure 3:**
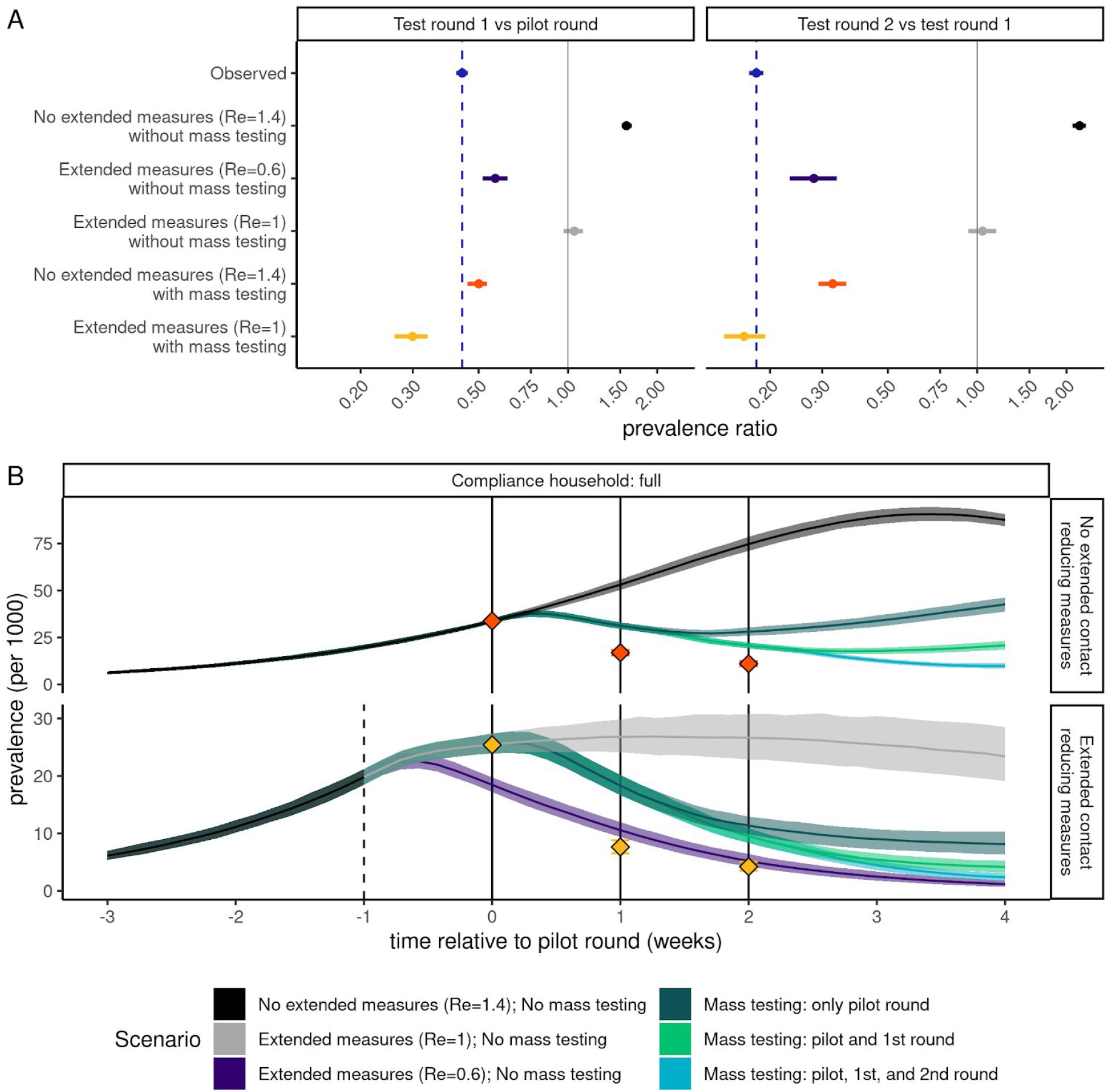
Simulated relative effectiveness of the extended contact reducing measures and the mass testing. Top panel: the change in observed prevalence of infectious non-quarantining individuals between 10 and 65 years of age as predicted by the microsimulation model. For comparison the observed test-positivity rate is shown in blue. The facets show changes from the pilot to the first round of mass testing (left) and from the pilot to the second round of mass testing (right). Shown scenarios compare the effect of (top to bottom) no additional interventions that limit the growth rate of R_e_=1.4, the extended contact reduction measures drastically reducing the growth rate to R_e_=0.6 and no mass testing being conducted, the extended contact reduction measures reducing the growth rate to R_e_=1.0 and no mass testing being conducted, no change in growth rate but mass testing, and the extended contact reduction measures reducing the growth rate to R_e_=1 and mass testing. In scenarios without mass testing, we compared prevalence of infectious individuals on the same days as testing occurred in scenarios with mass testing. Bottom panel: Simulated infection incidence of alternative intervention strategies. Simulations are aligned by the date of the first mass test (t=0). The dashed line indicates the timing of the extended contact reducing measures and the solid lines the timing of the mass testing campaigns. Colors indicate the simulations stratified into whether no mass testing or 1, 2 or 3 testing rounds were performed and the effectiveness of the extended contact reduction measures on the growth rate. Red and yellow dots indicate the prevalence of infectiousness observed among the tested non-quarantining age-eligible population, corresponding to the scenarios in the top panel.

**Table 1:**
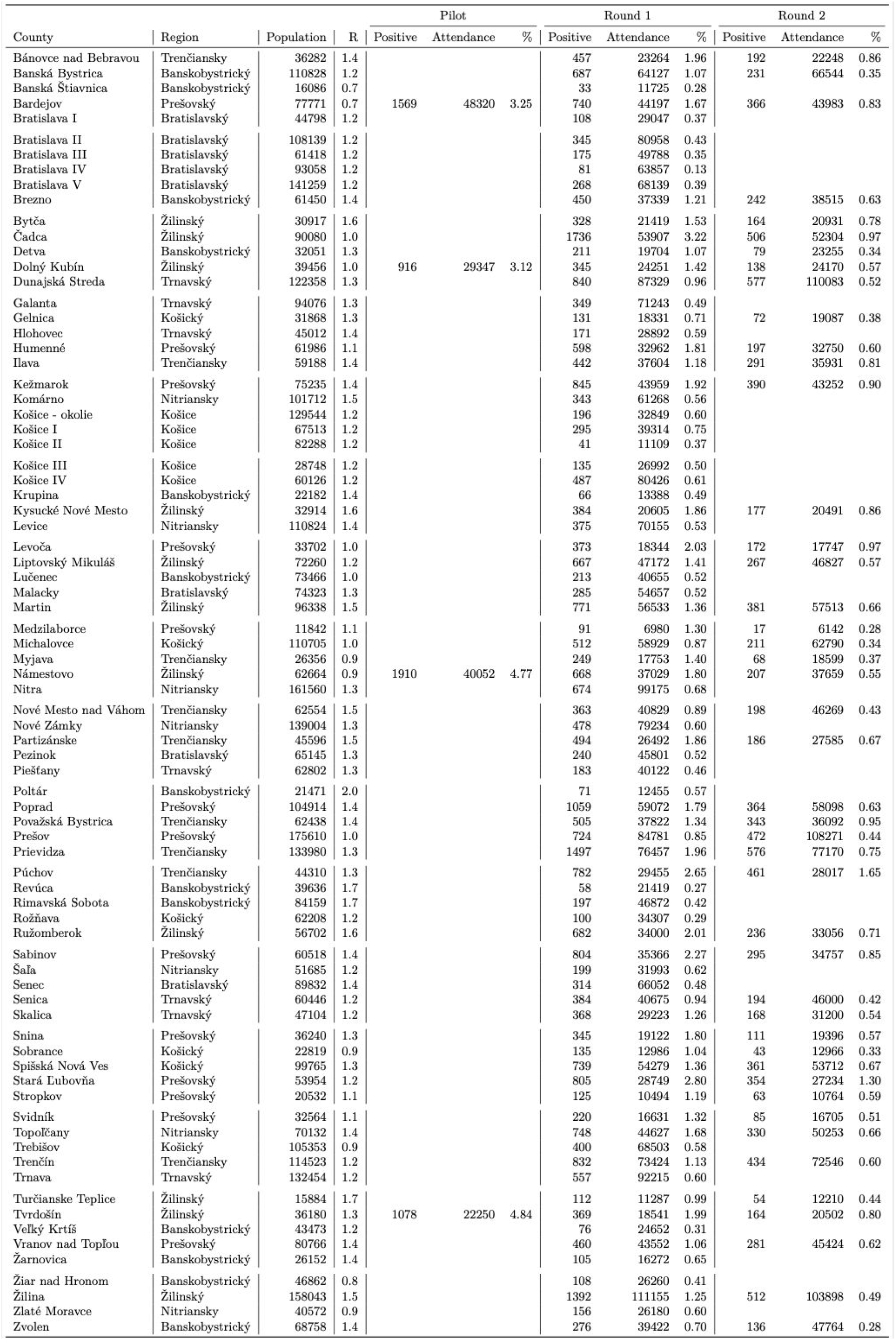
Overview of county specific test numbers and reductions for the 79 counties in Slovakia. R: median estimate of the reproduction number on 22 October, based on test-positive cases from syndromic surveillance up to 30 October and estimated using a renewal process model on back-calculated estimates of infection incidence. %: proportion positive out of those attending mass testing.

While we observed a reduction of more than 50% in test positivity between mass testing campaigns, the observed change in daily case incidence reported through standard surveillance was not the rapid collapse in test-positive cases that would correspond to the drastic reductions in prevalence. This may be due to a variety of reasons. Foremost, national mass testing campaigns are likely to have a major disruptive effect on passive syndromic surveillance. Also, the ability of PCR to detect viral RNA well beyond the infectious period will partially mask a sudden drop in infectious infections. In addition, starting mid-September the incidence surveillance has been operating at capacity with long waiting lists for testing and stricter eligibility criteria, which in the post mass testing period reduced substantially, and hence may have artificially reduced the observable change in such data. In contrast, data on hospital bed occupancy shows sudden flattening from mid-November suggesting a sharp decrease in new admissions consistent with a sizable reduction in new infections at the time of the mass testing campaigns (Figure S6).

Executing a large-scale mass testing campaign comes with several challenges. The need to mobilise sufficient medical personnel to conduct the nasopharyngeal swabs proved to be a major obstacle. Also, the logistics of mobilising large numbers of assisting army personnel and vast amounts of testing and PPE material proved challenging. Some of the challenges could be overcome by using other rapid antigen tests with similarly high sensitivity but which are also licensed for use with nasal swabs (*16, 17*). Nasal swabs can be self-administered and therefore reduce demand on trained personnel and transmission risk in the process of sample collection or even may enable testing at home. They are also less invasive and can be better suited for children and mass testing at schools. However, these benefits have to be weighed against the potential loss of sensitivity if self-administered (*18*), and the implications of the Slovak mass testing experience need to be studied carefully before considering potential replication elsewhere (*19*).

In conclusion, the combination of nationwide restrictions and mass testing with quarantining of household contacts of test positives rapidly reduced the prevalence of infectious residents in Slovakia. While impossible to disentangle the precise contribution of control measures and mass testing, the latter is likely to have had a substantial effect in curbing the pandemic in Slovakia and may provide a key tool in the containment of SARS-CoV-2.

## Material and Methods

### Study population

Slovakia is a country with a population of 5.5 million, consisting of 79 counties grouped into 8 administrative regions. Slovak residents aged between 10 and 65 years and older adults in employment were eligible for mass testing (about 4 million people). Those quarantining at the time or who had recovered from COVID-19 in the past three months were excluded.

The pilot was conducted in three counties in the Orava subregion (Námestovo, Tvrdošín, Dolný Kubín) and Bardejov county, which had the highest infection incidence detected in surveillance at the time. The first round of mass testing was conducted nation-wide and the second round of mass testing was restricted to 45 counties, mostly in the northern part of Slovakia, with observed infection prevalence in the first mass testing round exceeding 7 per 1,000 tests.

### Interventions

Slovakia implemented a series of infection control measures throughout October, which included closing schools for pupils aged 14 or above on 15 Oct and for pupils aged 10 and above on 26 October. They remained closed throughout the period of the mass testing campaigns and thereafter. Indoor gastronomy and indoor leisure activities were also restricted. Residents were further asked to limit their movement for one week between 24 October and 1 November only to: going to work, taking children to school, shopping for essential items and going for recreational walks (Figures 1 and S4). Although these rules were legally enforceable, Slovakia relied mostly on people’s civil responsibility to adhere to restrictions.

On the days of mass testing, participants attended testing centres run by healthcare professionals, armed forces and volunteers. Overall, Slovakia deployed around twenty thousand medical staff and forty thousand non-medical personnel. Testing procedures followed as recommended by the manufacturer, with nasopharyngeal samples obtained by trained medical personnel using flexible, aluminum-shaft, calcium alginate swabs (*20*).

Testing was not obligatory, but residents who did not attend the mass testing were instructed to stay home for ten days or until the next round of mass testing. A medical certificate was issued to every participant confirming their infection status. A test-negative certificate was required by employers to enter workplaces. Various venues and public institutions inspected peoples’ certificates at random. Private PCR tests were also accepted if no older than the most recent mass testing campaign. Citizens whose test results were positive were asked to enter a 10-day long quarantine together with all members of the same household and their self-traced contacts in the preceding two days in an attempt to reduce secondary transmission. While both household and contact quarantine were strongly encouraged, no control or enforcement measures were in place. Individuals currently in quarantine were asked not to attend the mass testing.

### Data

No participant information was collected during either of the mass-testing campaigns. Hence, none of the analyses could be stratified by age or other demographic factors. However, information on the number of tests used as well as the number of positive tests has been tracked and made openly available by the Slovak Government (*12*). The SD Biosensor Standard Q antigen test that was used exclusively has high specificity, with point estimates typically in excess of 99.5%. Sensitivity exceeded 70% in most validation studies, and exceeded 90% among samples with a low cycle threshold, which correlates with effective transmission (*13, 21, 22*). For example, a recent validation study in the UK estimated the sensitivity of lateral flow devices in detecting infectious individuals to be in the order of 83-91% (*11*).

To assess trends in the local epidemiology of SARS-CoV we used routine syndromic and PCR confirmed surveillance for the daily incidence of infections as reported by the Slovak Ministry of Health (*23*).

### Analyses

We explored the relationship between county level prevalence as measured in the first round of mass testing and a range of demographic (mean age, population density), socioeconomic (unemployment rate), epidemiological (inclusion in the pilot testing) and ethnic (size of the Roma population) predictors in a Bayesian hierarchical negative binomial regression.

We estimated the impact of mass testing via the change in test positivity rates in counties with at least two mass testing campaigns. Test positivity rates in the study’s context are an estimate for the prevalence of infectious SARS-CoV-2 infections (unaccounted for the sensitivity and specificity of the test). We calculated crude prevalence ratios (cPR) to estimate the change in test positivity between mass testing campaigns, including Wald-Normal confidence intervals. Binomial confidence intervals were calculated for the proportion of tests that were found positive. Test positive rates provide a natural upper bound for false positive rates of a test. We thus estimated the minimum test specificity *ms* as the probability of observing a test positivity of at least 1-*ms* in at least one county, assuming the test positivity to be binomially distributed. Test sensitivity could not be estimated in this study.

To explore heterogeneity between counties in the estimated reduction in test positivity in subsequent rounds of mass testing, we used a quasi-Poisson regression model. The number of positive tests in each county was modelled with a county specific intercept, an indicator variable for the round 2 of mass testing, and interactions of the latter with attendance rates in round 1, round 1 test positivity, the reproduction number leading up to round 1 and region as covariates as well as the log number of tests as an offset variable. The three continuous variable interaction terms were centered and standardised (see supplement).

We used the EpiNow2 model (*24*) for the calculation of trends in local epidemiology prior to mass testing based on routinely reported infection incidence. EpiNow2 uses observed delay distributions in combination with a renewal equation model to probabilistically infer the infection date for each reported case as well as the population-wide time varying reproduction number, allowing a smoothed extrapolation of infection incidence and prevalence and extrapolation beyond the observed study period under an assumption of no change. We define the self-adjusted prevalence ratio (saPR) as the cPR divided by the prevalence ratio at the times of round 2 vs round 1 as estimated through EpiNow2. The saPR is an estimate for the effect of the intervention that takes into account that infection prevalence would have changed in the time between observations (see supplement).

To explore scenarios for the relative effect of mass testing and extended contact reducing measures we used a microsimulation model. We focused on three scenarios in which mass testing takes place, i) an epidemic growth rate of R_e_=1.4 (as in early October) that is unchanged by any additional contact reducing measures measures, ii) a reduced growth rate of R_e_=1 from 15^th^ October (similar to many parts of Europe in the weeks following autumn lockdowns) and iii) the growth rate reduced to R_e_=0.6 from 15^th^ October (the smallest observed reproduction number nationally during the COVID-19 pandemic) but no effect of mass testing. A detailed model description is provided in the supplementary material, but in brief: Individuals are grouped in households according to Slovak census data (*25*), and make contact with individuals outside their household at age-specific rates (*26*). To account for social distance measures, we assumed absence of at-school contacts for children 10 years and over, and that contacts at work and contacts not at the home, school, or workplace, were reduced by 25% and 75% from pre-epidemic levels, respectively. We simulated infections among 78,000 susceptible individuals, representative of the population size of a typical pilot county. When infection prevalence reached 3.2% (approximating a typical observed prevalence during the testing pilot), up to 3 rounds of weekly mass testing were initiated and the week before that restrictions equivalent to those enacted in Slovakia were implemented. In the model, we assumed perfect test sensitivity for detection of currently infectious infections, specificity, and compliance with quarantine. Observed test attendance rates were used assuming that individuals in quarantine did not attend mass-testing.

### Open Access

Daily incidence of positive COVID-19 test reports and the results of the mass testing are available through governmental websites (*12, 23*). All analyses were conducted in R (*27*) and can be found at www.github.com/sbfnk/covid19.slovakia.mass.testing (data analyses) and https://github.com/kevinvzandvoort/covid_svk (simulation model).

## Data Availability

Daily incidence of positive COVID-19 test reports and the results of the mass testing are available through governmental websites and included in the code repository. All analyses were conducted in R and can be found at www.github.com/sbfnk/covid19.slovakia.mass.testing (data analyses) and https://github.com/kevinvzandvoort/covid_svk (simulation model).

## Acknowledgements

We would like to thank all healthcare workers, Slovak armed forces and countless volunteers who helped with the execution of the mass testing campaign. Lastly we would like to thank all participants who contributed their time to help curb the Pandemic and particularly those who had to quarantine as a result of their or their household’s or contact’s test result.

The following authors were part of the Centre for Mathematical Modelling of Infectious Disease COVID-19 Working Group. Each contributed in processing, cleaning and interpretation of data, interpreted findings, contributed to the manuscript, and approved the work for publication: Gwenan M Knight, Naomi R Waterlow, Carl A B Pearson, Fiona Yueqian Sun, Simon R Procter, Alicia Showering, Rosalind M Eggo, Yung-Wai Desmond Chan, Emily S Nightingale, David Simons, Oliver Brady, Billy J Quilty, Petra Klepac, Amy Gimma, Hamish P Gibbs, W John Edmunds, Adam J Kucharski, Sam Abbott, Jack Williams, Kiesha Prem, Rosanna C Barnard, Thibaut Jombart, Graham Medley, Katherine E. Atkins, Samuel Clifford, Nicholas G. Davies, Kaja Abbas, Mark Jit, Timothy W Russell, Frank G Sandmann, Damien C Tully, James D Munday, Anna M Foss, Alicia Rosello, Sophie R Meakin, Joel Hellewell, C Julian Villabona-Arenas, Christopher I Jarvis, Rachel Lowe, Akira Endo, Matthew Quaife, Nikos I Bosse, Yang Liu.

This work is licensed under a Creative Commons Attribution 4.0 International (CC BY 4.0) license, which permits unrestricted use, distribution, and reproduction in any medium, provided the original work is properly cited. To view a copy of this license, visit https://creativecommons.org/licenses/by/4.0/. This license does not apply to figures/photos/artwork or other content included in the article that is credited to a third party; obtain authorization from the rights holder before using such material.

## Supplementary material to “The effectiveness of population-wide screening in reducing SARS-CoV-2 infection prevalence in Slovakia”

### Supplementary Tables and Figures

**Table S1:** Regression model coefficients. These are the fitted values for the negative binomial regression model used to estimate the adjusted prevalence ratio (aPR = exp(*δ*_*1*_)). The associated prevalence ratios are visualised in supplementary Figure S8.

| Variable | Lower 95% | Median | Upper 95% |
| --- | --- | --- | --- |
| Intercept | 0.0084 | 0.0098 | 0.011 |
| Round | 0.41 | 0.44 | 0.48 |
| Mean age | 0.61 | 0.73 | 0.87 |
| Population density | 0.73 | 0.82 | 0.93 |
| Unemployment rate | 0.72 | 0.92 | 1.2 |
| Roma population | 0.69 | 0.88 | 1.1 |
| Previous attendance | 0.86 | 0.99 | 1.1 |
| Previous prevalence | 0.88 | 0.95 | 1 |
| Reproduction number | 0.99 | 1.1 | 1.2 |

**Figure S1:**
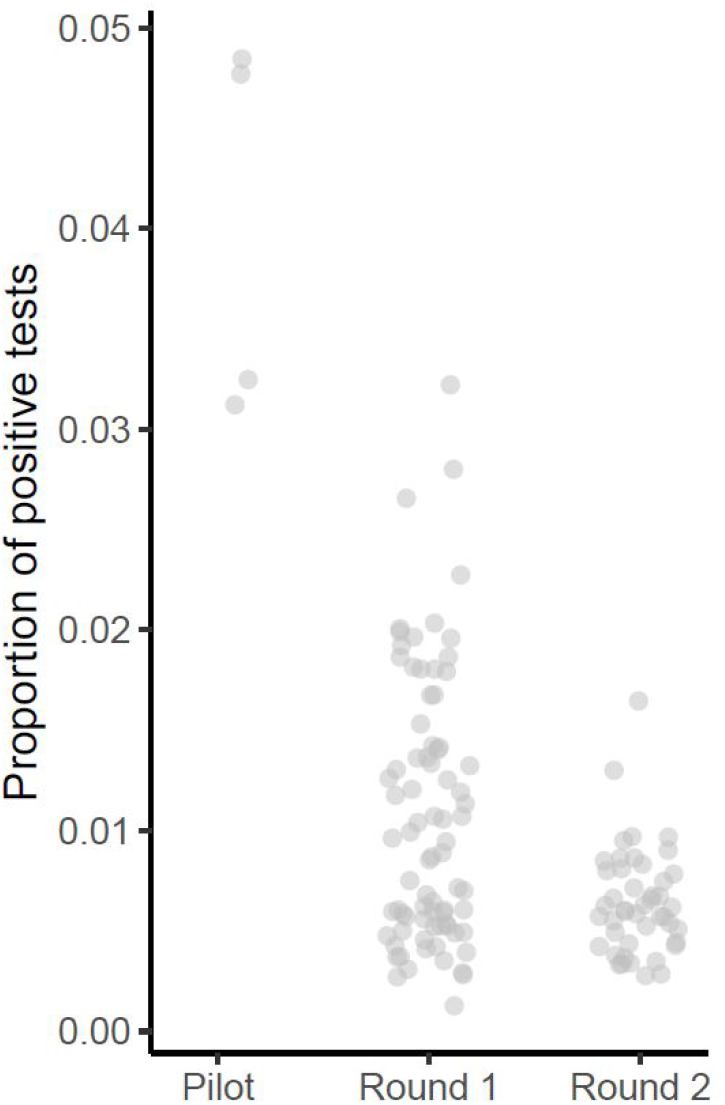
Proportion of positive tests. Test positivity grouped by different mass testing rounds. Given a sufficiently large sample size, one minus test specificity would be the lowest observable proportion of positive test. The absence of apparent clustering of observations at the lower end of the observed range suggests that even lower value could have been observed and test specificity was not a limiting factor.

**Figure S2:**
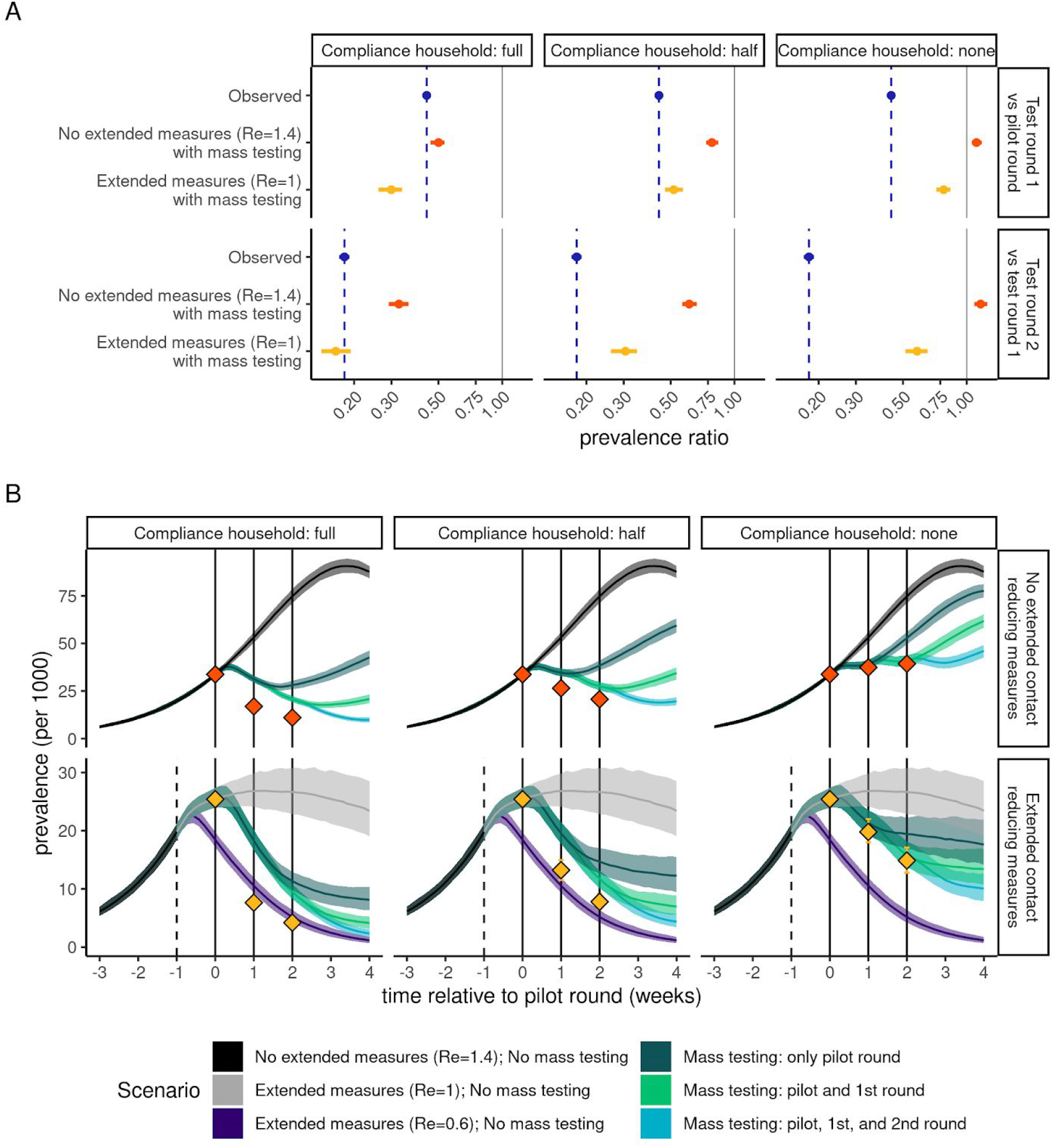
Simulated relative effectiveness of the extended contact reducing measures and the mass testing with different levels of adherence to quarantine for household members of test-positives. Top panel: the change in observed prevalence of tested infectious non-quarantining individuals between 10 and 65 years of age as predicted by the microsimulation model. For comparison the observed test-positivity rate is shown in blue. The facets show changes from the pilot to the first round of mass testing (top) and from the pilot to the second round of mass testing (bottom). The horizontal facets show different levels of adherence to quarantine for household members of test-positives, from full adherence (100% compliance; left), half (50% compliance, middle), to no (0% compliance, right). Shown scenarios compare the effect of mass-testing without (orange) and with (yellow) additional contact reducing measures. Bottom panel: Simulated infection incidence of alternative intervention strategies with different levels of adherence to quarantine for household members of test-positives (in horizontal facets). Simulations are aligned by the date of the first mass test (t=0). The dashed vertical line indicates the timing of the extended contact reducing measures and the solid vertical lines the timing of the mass testing campaigns. Colors indicate the simulations stratified into whether no mass testing or 1, 2 or 3 testing rounds were performed and the effectiveness of the contact reducing measures on the growth rate. Red and yellow dots indicate the prevalence of infectiousness observed among the non-quarantining age-eligible population, corresponding to the scenarios in the top panel under the same level of compliance with quarantine for household members of test-positives.

**Figure S3:**
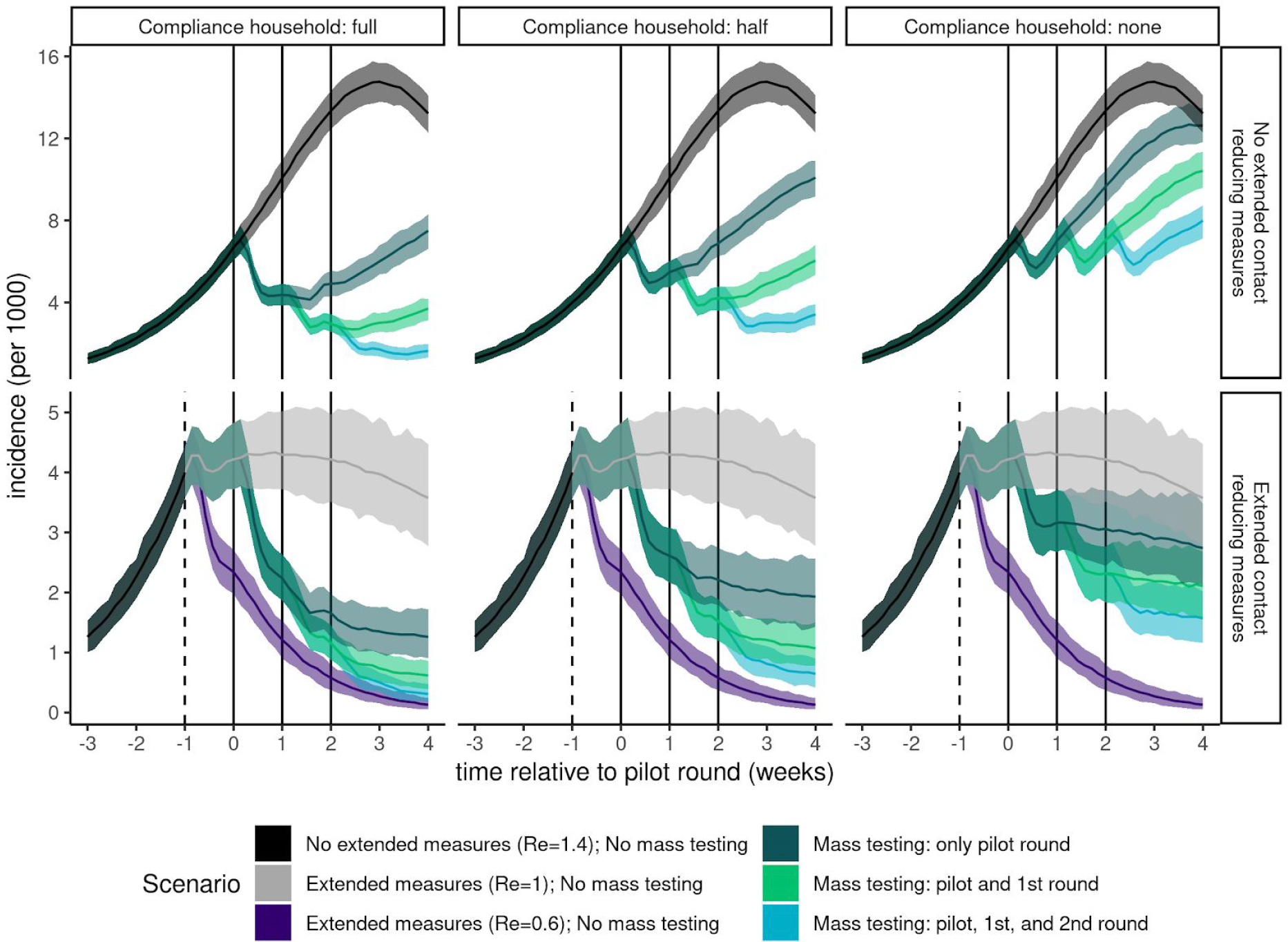
Simulated impact of the extended contact reducing measures and the mass testing over time on incidence with different levels of adherence to quarantine for household members of test-positives. Simulated daily infection incidence of alternative intervention strategies. Simulations are aligned by the date of the first mass test (t=0). The dashed line indicates the timing of the extended contact reducing measures and the solid lines the timing of the mass testing campaigns. Colors indicate the simulations stratified into whether no mass testing or 1, 2 or 3 testing rounds were performed. In the full household compliance facets all household members quarantine for 10 days if a member was tested positive and in the non compliance facet they did not.

**Figure S4:**
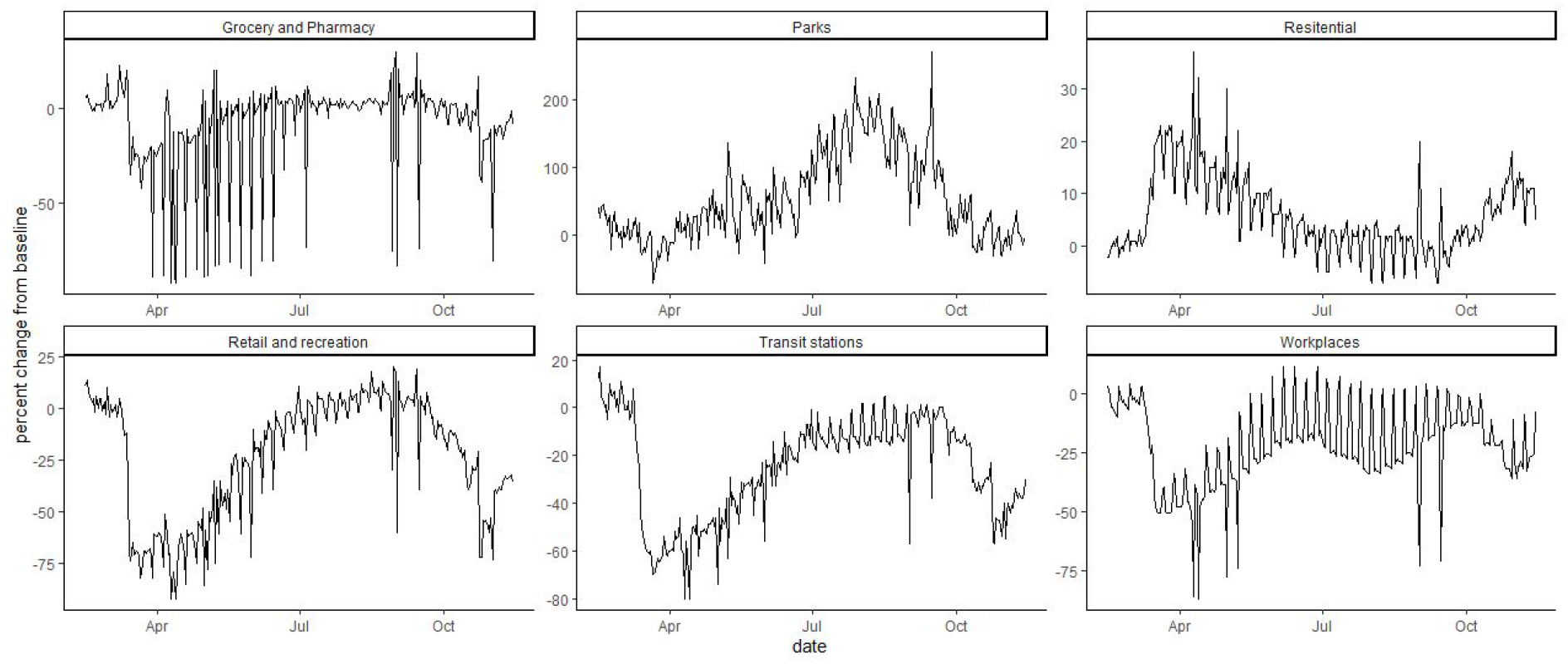
Google mobility index for Slovakia. The change in mobility in comparison to baseline for a number of settings during 2020 in Slovakia. The mobility data is as provided by Google (https://www.google.com/covid19/mobility/).

**Figure S5:**
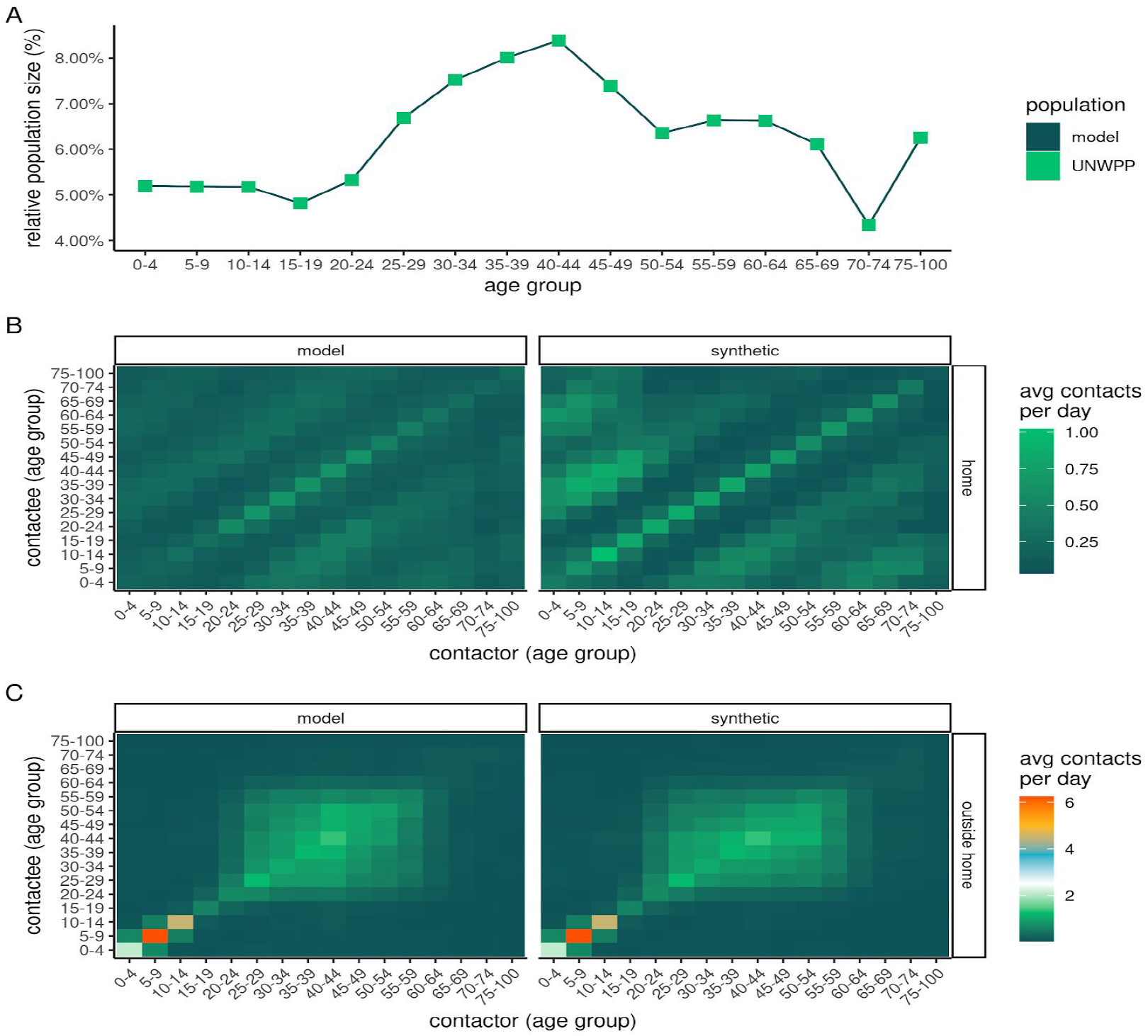
Comparing the microsimulation model population to observed structures in Slovakia. Panel A shows the median relative population distribution across all model runs (dark-green) compared to the UNWPP population estimates for Slovakia in 2020 (light-green), by age-group. Panel B shows the median household contact matrix (left; assuming all household members make one contact per day) compared to the synthetic household contact matrix (right), adjusted for UNWPP population size. Panel C shows the median non-household contact matrix (left) compared to the synthetic non-household contact matrix (right), adjusted for extended contact reducing measures and UNWPP population size.

**Figure S6:**
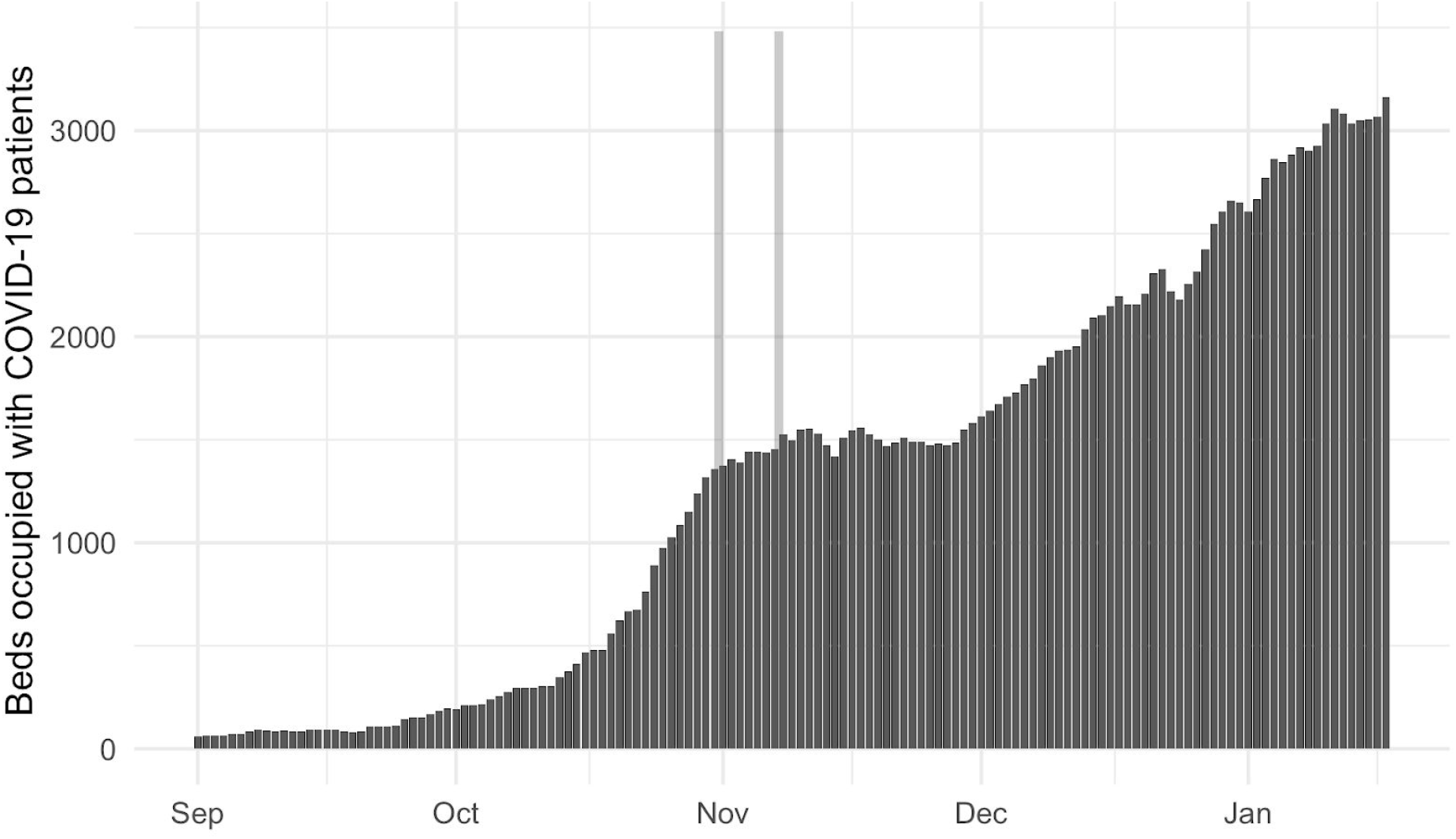
Daily hospital bed occupancy with COVID-19 patients in Slovakia during the autumn of 2020. Following an increase particularly during October a sharp the abrupt levelling off in the first week of November suggests a sharp decrease in new admissions following the mass testing (dates of round 1 and 2 shown as grey bars). Data presented are available from the European Centre for Disease Prevention and Control (https://www.ecdc.europa.eu/en/publications-data/download-data-hospital-and-icu-admission-rates-and-current-occupancy-covid-19)

**Figure S7:**
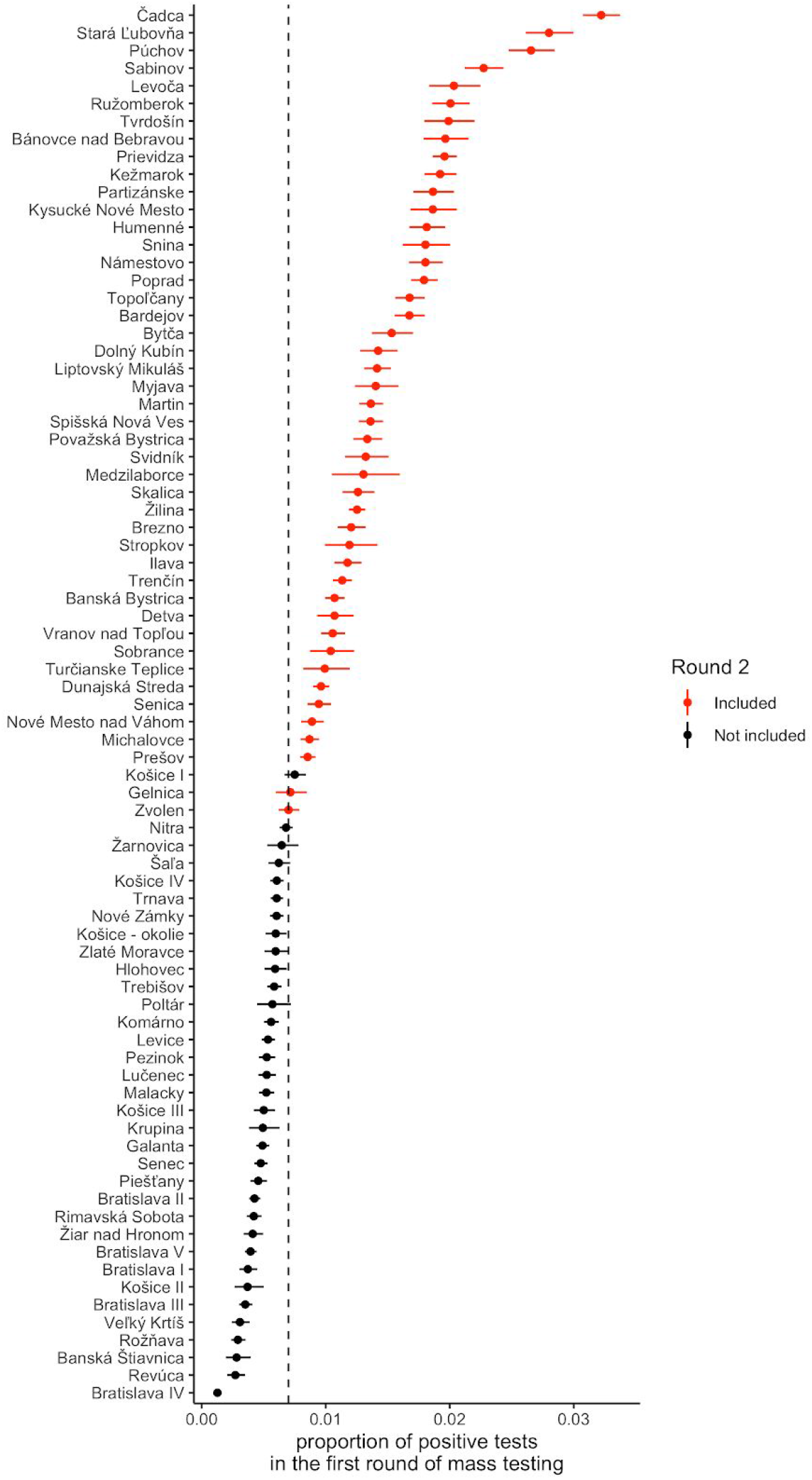
Test positivity rates for each county in the first round of mass testing. Between county heterogeneity in test positivity in the first mass testing campaign in Slovakia. Point estimates are displayed as dots and binomial confidence intervals as lines. The dotted line indicates the test-positivity threshold used for determining which counties were included into a second national mass testing round, coloured in red.

**Figure S8:**
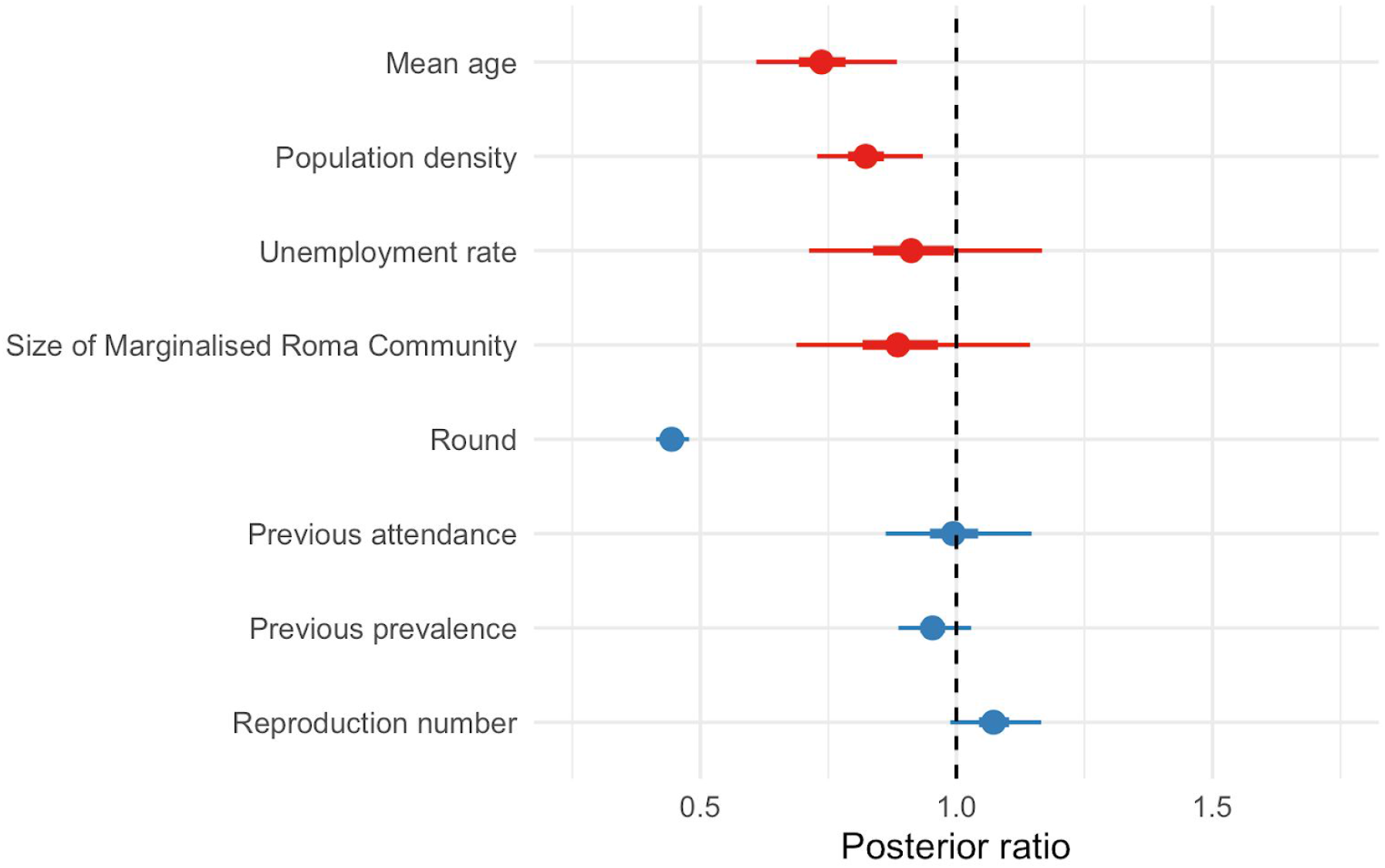
Estimated association of possible predictors with prevalence of test-positivity at the county level. Coefficients were determined in negative binomial regression (size: total number tested) with the number testing positive in the first round of mass testing in each county (pilot or nationwide testing) modelled as a function of standardised variables representing mean age, population density, unemployment rate and size of the Marginalised Roma Community (red), and the number testing positive in subsequent rounds additionally as a function of a mass testing effect (“Round”), attendance and prevalence in the previous round, and the reproduction number on 22 October. Shown are the median estimated posterior ratios with 50% and 95% credible intervals, i.e. the exponentiated coefficients from Supplementary Table S1 which in the case of the *Round* coefficient corresponds to the adjusted prevalence ratio (aPR) associated with mass testing.

### Additional details for the study

Detailed timeline of national SARS-CoV-2infection control measures adopted in Slovakia

#### Pre - 1 October

- Compulsory face coverings indoors, in enclosed public places and inside mass transport vehicles
- 1000 limit on number of people in aquaparks
- 1000 outdoors and 500 indoors limit on mass gatherings
- Travellers returning from “high risk” countries or regions are requested to take a PCR test after the fifth day of their arrival or remain in quarantine for 10 days
- Shopping hours between 9am and 11am reserved for the elderly

#### 1 October

- Gatherings limited to max 50 people
- Wedding receptions banned

#### 15 October

- Gatherings limited to max 6 people (indoors or outdoors)
- Online schooling for pupils aged 14 years or older
- Compulsory face coverings including outdoors, if within city limits
- Wake receptions banned
- Indoor gastronomy closed
- Theatres and cinemas closed
- Pubs, clubs and bars closed
- Gyms, swimming pools, aquaparks, spas and other wellness and fitness facilities closed
- Church and religious services suspended

#### 24 October - 1 November

- National stay at home order (lockdown) with the following exceptions:
  - travel to and from place of work
  - accompanying children to and from school
  - the first four grades of elementary schools, nurseries and creche stayed open
  - essential travel and activities (i.e. groceries, pharmacy, doctor surgeries, caring for a family dependant, animal husbandry, walking pets within 100 meter distance from home, funerals, post office, bank, insurance company, cleaning services, car repair services, petrol stations)
  - recreational nature walks

#### 2 November

- same restrictions as 15 October with the addition of closing school for pupils aged 10 year or older.

##### EpiNow2

We used EpiNow2 to backcalculate infection curves in pilot and non-pilot regions. These were converted to infection prevalence using a detection window of 2-6 days after exposure. This allowed us to estimate the infection prevalence of reported cases at the time of mass testing (*p1*) and in the subsequent mass testing round (*p2*). Thus we define the self adjusted prevalence ratio as the crude prevalence ratio observed in the mass testing campaigns adjusted for the predicted change in prevalence if no mass testing or other interventions were conducted:

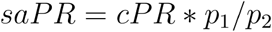

#### Model for round 1 prevalence

##### Regression model

We used a negative Binomial regression model that was a priori defined by a choice of available covariates that could have plausibly explained the observed prevalence and the impact of the intervention. The observed number of positive tests in country *i* during round *r*

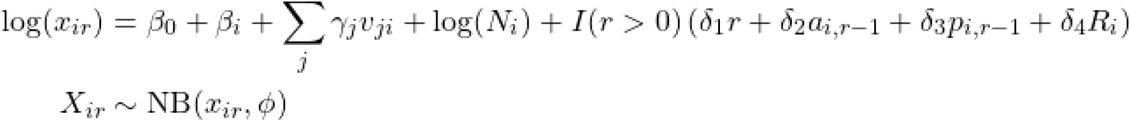

where I(r > 0) is an indicator function that is 1 if r > 0 and 0 otherwise, and

*i* = county indicator

*r* = round indicator: 0 for the first round of mass testing in the county (pilot in the countries selected for the pilot, nationwide testing in all other counties), increased by one for each subsequent round

*β*_*0*_ = global intercept

*β*_*i*_ = group-level intercept in for county *i*

*γ*_*j*_ = coefficient of prevalence covariate *v_ij_*

*v*_*ij*_ = centered and standardised value of covariate *j* for prevalence in county *i* (where *j* is one of: mean age, population density, unemployment rate and size of the Marginalised Roma Community)

*δ*_*j*_ = coefficient of prevalence reduction covariate *j (r, a*_*i,r*_, *p*_*i,r*_ and *R*_*i*_)

*N*_*i*_ = number of people tested in each county in mass testing rounds 1 and 2

*a*_*i,r*_ = attendance rate of mass testing round *r* in county *i* as a proportion of the total population

*p*_*i,r*_ = prevalence observed in mass testing round *r* in county *i*

*R*_*i*_ = net reproduction number estimated from EpiNow2 on 22 October in county *i*

The adjusted prevalence ratio (aPR) between rounds can then be calculated as 1-exp(*δ*_*1*_).

#### Microsimulation model

##### Model structure

We used an individual-based, probabilistic microsimulation model (IBM) to study the expected reduction in prevalence of (detected) infectiousness under different assumptions.

We parameterized our model to represent an average county of Slovakia.

In our IBM, individuals fall within *a* age strata (where *i* is a given age stratum) with relative proportions *p*_*i*_. They belong to households of mean size *m*_*h*_ (we combine different datasets to simulate a population). The simulation starts when the model population of size *N* days. is seeded with at least one SARS-CoV-2 infection, and runs for 365

Births, non-COVID-19 deaths, ageing and migration are omitted from the model given its short timeframe. The study’s endpoint of interest is infection, we did not include hospitalisation or clinical outcome status of cases. Infectiousness is assumed to be unaffected by clinical severity, but does differ for asymptomatic, pre-symptomatic and symptomatic cases (see below).

##### Infection states and transitions

At any time *t*, individuals within the IBM are within one of the following classes: *S* (susceptible), *E* (exposed and latent, i.e. infected but not yet infectious), *I*_*P*_ (infectious but pre-symptomatic), *I*_*C*_ (infectious and symptomatic), *I*_*S*_ (infectious and asymptomatic throughout the infection), or *R* (removed: recovered and assumed to be immune or deceased). The age-specific probability of becoming a symptomatic case when infected is *y*_*i*_

Over any Δ*t* time unit, any given individual has the following binomial probabilities of transitioning to a subsequent state:

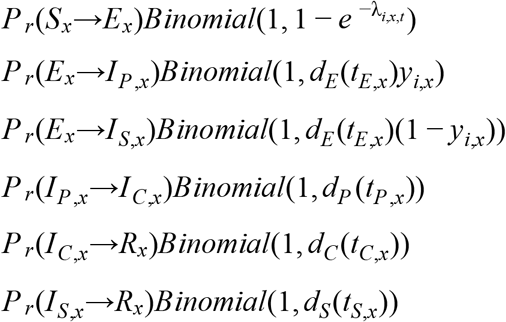

where 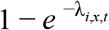 is the age-specific instantaneous force of infection experienced by a susceptible individual, as detailed below; and, *d*_*E*_, *d*_*P*_, *d*_*C*_, and *d*_*S*_ are cumulative distribution functions (CDFs) for the duration of the corresponding states: *d*_*E*_ (*t*_*E,x*_) denotes the CDF for the duration of the pre-infectious state evaluated at the time already spent by individual *x* in that state, and so on.

##### Transmission dynamics

Over any Δ*t* time unit, susceptible individuals of any age *i* within each household *h* move from *S* to *E* based on an individual-specific instantaneous force of infection that is the sum of λ due to contacts within the household and λ due to extra-household contacts:

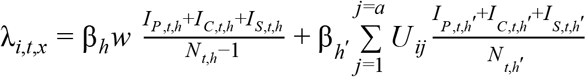

where β_*h*_ and β_*h*_′ are the probabilities of transmission per contact between a susceptible and infectious person for contacts made within and outside the household itself, *f* is the relative infectiousness of asymptomatic infections, compared to cases that do develop symptoms, *w* is the mean per-capita intra-household contact rate, assuming random mixing within the household. *U* is the contact matrix outside the household for the total number of contacts made between individuals aged i with individuals aged j. *h* denotes individuals within the household itself, while *h*^′^ denotes individuals in the population excluding the household itself). *I*_*P, t*_, *I*_*C,t*_, and *I*_*S,t*_ represent the total number of infectious individuals not in quarantine at time *t*.

We assume that all individuals within the household contact each other once per day, and calculate the expected population-wide intra-household contact matrix *W* where *W* _*ij*_ is the sum of all aged individuals aged *i* living together with household members of age *j*, divided by the model population size aged *i*. We ensure that the average contact rates are such that the total number of extra-household contacts are symmetric between age-groups, and calculate the population-level contact matrix, *Z* = *W* + *U*.

The basic reproduction number *R*_0_ is then defined as the average number of secondary infections generated by a typical infected individual in a fully susceptible population not affected by any contact reducing measures (where β_*h*_′ = β_*h*_ = β), and is computed as the dominant eigenvalue of the next generation matrix (NGM) of the corresponding compartmental model structure to our IBM model, defined as:

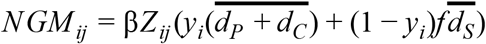

where accents indicate the expected (average) values. Lastly, β is the ratio of this eigenvalue and the *R*_0_ value assumed in the simulation (see below).

We validated the calculated *R*_0_ value through this method by running multiple iterations of the model using a different seed for the random number generator, and calculating the average number of secondary cases derived from all infectious individuals who completed their period of infectiousness in the first 30 days of the simulation.

##### Mass-Testing and extended contact reducing measures

We simulate an epidemic using a timestep of Δ*t* = 1*day*. The pilot round of mass testing is introduced at time *t*_*g*_ when the prevalence of infectiousness in the model reaches a predefined threshold (average of observed prevalence in the pilot counties). In scenarios in which additional rounds of mass testing are introduced, these first and second rounds of nationwide mass testing are introduced on days *t*_*g*_ + 7 and *t*_*g*_ + 14.

When testing is introduced, we assume that any individual *x* attends mass-testing with probability *z*_*t*_. We calculated this probability as 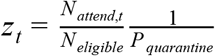, where *N*_*attend,t*_ is the observed attendance for the test round introduced at time *t, N*_*eligible*_ is the total model population size that is eligible for testing (any individual between the ages of 10 and 65), and *P*_*quarantine*_ is the proportion of the model population size that is in quarantine at time *t*.

Individuals already in quarantine do not attend testing. We assume 100% sensitivity to detect an infectious individual (in state *I*_*P*_, *I*_*S*_, or *I*_*C*_), 0% sensitivity to detect an infected but not yet infectious individual (in state *E*), and 100% specificity for any individual not currently infected (a full list of assumptions, including their sources, are listed in Table S2). Those who test positive are assumed to comply with quarantine measures with probability *C*_*p*_, and any of their household members not already quarantining are assumed to comply with probability *C*_*h*_. We also assume the same probability *C*_*h*_ to quarantine individuals who do not attend mass-testing, but are eligible (between the ages of 10 and 65).

To implement scenarios with extended contact reducing measures, we first calculated the effective reproduction number in the two weeks before the first round of mass-testing would be implemented, between *t*_*g*_ − 14 and *t*_*g*_. We then started a new model run using the same seed for the random number generator, and implemented extended contact reducing measures that affect community transmission but not intra-household transmission by changing the value for the probability of effective contacts made outside of the household β_*h*_′ from the time of implementation of these measures (at *t*_*g*_ − 7) with 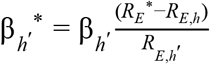, where *R*_*E,h*_ and *R*_*E,h*_′ are the estimated effective reproductive numbers for transmission within and outside the household in the period before implementation of contact reducing measures, and *R*_*E*_^*^ is the target value for the effective reproduction number after implementation of contact reducing measures. We assumed the reduced β_*h*_′^*^ would remain in place for the remainder of the simulation.

##### Population structure

We simulate a new population within each model iteration by combining estimates for the 2020 Slovak population size, household size by age, and the estimated number of daily contacts made in the household per day.

We simulated a population with target size *N* by simulating new households until the sum of individuals in all households reached *N*.

To simulate a household, we randomly sampled *i*, the age of one individual living in the new household, and drew a value for *Y*, the household size (ranging from 1 to 6) for those living in the household, from a multinomial distribution where the age-specific distribution of household sizes as estimated in the 2011 Slovak census were used as probabilities for the household size (Eurostat, 2020).

We assumed that normalized age-specific at-home contact rates, *W* _*ij*_ ^*^, calculated as 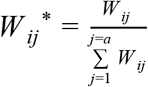, were proportional to household age distribution (*26*).

We then sampled *Y* – 1 age-groups of household members from a multinomial distribution with age-specific probability of sampling age-group *j, P* (*j*|*i*) = *W* _*ij*_^*^*p*_*j*_, where *P* _*j*_ is the probability of sampling any individual from age-group *j*, following age-specific United Nations World Population Prospects (UNWPP) estimates for the population size (UNWPP, 2019).

The median average household size across all modelled populations is 3.7 (3.6-3.7). This is slightly lower than the average household size across all age groups (4.0) as reported in the 2011 Slovak household census (2020, Eurostat - Population by sex, age group, size of household and NUTS 3 regions). Figure S5 compares other key model parameters for the simulated populations with the empirical datasets used. Panel A compares the UNWPP population distribution for Slovakija in 2020 with the median population distribution across all simulated populations. A black area underneath the median population size shows the 95% interval of estimates across all populations, but is not visible in the plot as there is barely any variability across simulated populations, due to the algorithm that was used.

Panel B compares the median household contact matrix across all simulated populations to the synthetic at home contact matrix, where the synthetic matrix has been adjusted with the UNWPP population size estimates to ensure symmetry in the total number of contacts (i.e. total number of contacts of those aged i with j = total number of contacts of those aged j with i). We used the dominant eigenvalue of all matrices to select the matrix representing the median model matrix. The matrices are very similar, though there are slightly less child-adult contacts in the median model matrix compared to the synthetic matrix. The synthetic matrix is generated through extrapolation of contact surveys done in the mid 2000s in other European countries, and may therefore not reflect actual household contact patterns in Slovakia. In addition, the surplus of contacts in the synthetic contact matrix could be due to inclusion of extra-household contacts occurring at the home, which are not included in the model household contact matrix.

Panel C compares the median contact matrix for contacts made outside of the household used in the model, with the contact matrix for non-home contacts in the synthetic matrix for Slovakija (adjusted to represent a change in contact patterns due to Covid-19 interventions). The model contact matrices have been made symmetric for the population distribution used in the model, while the synthetic contact matrix has been made symmetrical for the UNWPP 2020 Slovakija contact matrix, but are otherwise identical. As these population distributions are very similar (Panel A), the contact matrices are as well.

##### Parameter values

The table below lists all parameter values used in the model

**Table S2:**
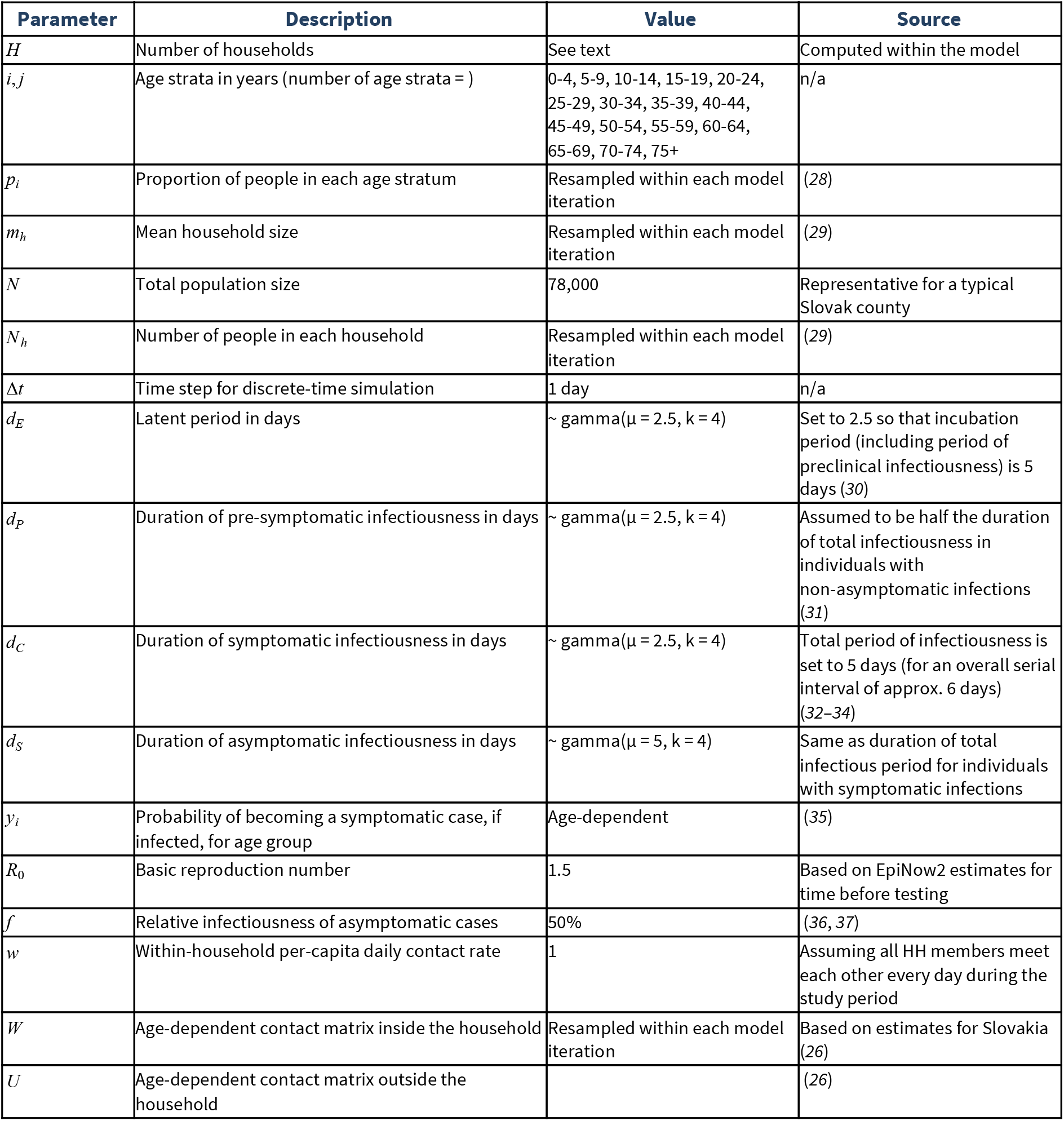
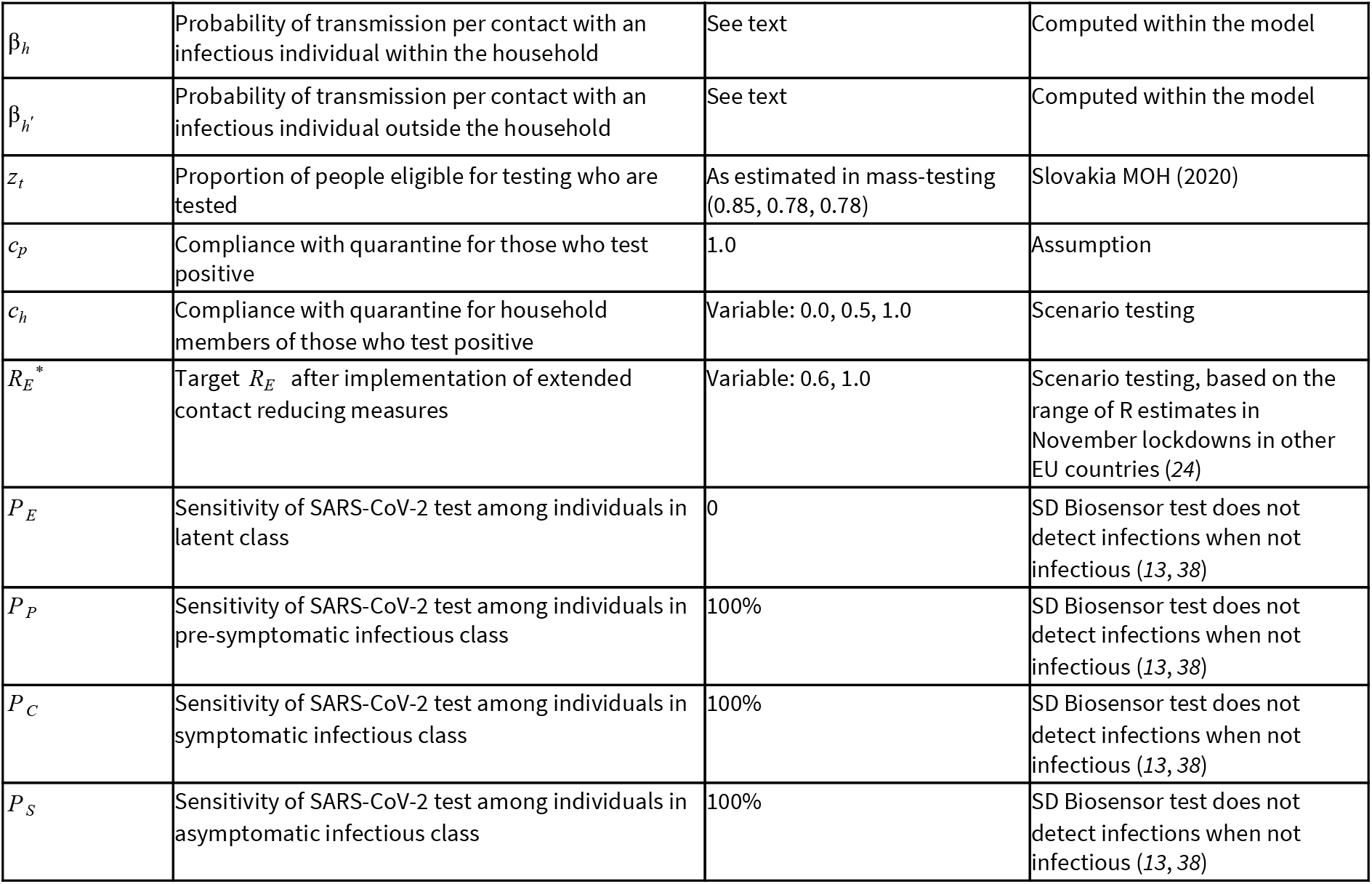
Parameter values used in the model.

| Parameter | Description | Value | Source |
| --- | --- | --- | --- |
| $H$ | Number of households | See text | Computed within the model |
| $i, j$ | Age strata in years (number of age strata = ) | 0-4, 5-9, 10-14, 15-19, 20-24, 25-29, 30-34, 35-39, 40-44, 45-49, 50-54, 55-59, 60-64, 65-69, 70-74, 75+ | n/a |
| $p_i$ | Proportion of people in each age stratum | Resampled within each model iteration | (28) |
| $m_h$ | Mean household size | Resampled within each model iteration | (29) |
| $N$ | Total population size | 78,000 | Representative for a typical Slovak county |
| $N_h$ | Number of people in each household | Resampled within each model iteration | (29) |
| $\Delta t$ | Time step for discrete-time simulation | 1 day | n/a |
| $d_E$ | Latent period in days | $\sim \text{gamma}(\mu = 2.5, k = 4)$ | Set to 2.5 so that incubation period (including period of preclinical infectiousness) is 5 days (30) |
| $d_P$ | Duration of pre-symptomatic infectiousness in days | $\sim \text{gamma}(\mu = 2.5, k = 4)$ | Assumed to be half the duration of total infectiousness in individuals with non-asymptomatic infections (31) |
| $d_C$ | Duration of symptomatic infectiousness in days | $\sim \text{gamma}(\mu = 2.5, k = 4)$ | Total period of infectiousness is set to 5 days (for an overall serial interval of approx. 6 days) (32–34) |
| $d_S$ | Duration of asymptomatic infectiousness in days | $\sim \text{gamma}(\mu = 5, k = 4)$ | Same as duration of total infectious period for individuals with symptomatic infections |
| $y_i$ | Probability of becoming a symptomatic case, if infected, for age group | Age-dependent | (35) |
| $R_0$ | Basic reproduction number | 1.5 | Based on EpiNow2 estimates for time before testing |
| $f$ | Relative infectiousness of asymptomatic cases | 50% | (36, 37) |
| $w$ | Within-household per-capita daily contact rate | 1 | Assuming all HH members meet each other every day during the study period |
| $W$ | Age-dependent contact matrix inside the household | Resampled within each model iteration | Based on estimates for Slovakia (26) |
| $U$ | Age-dependent contact matrix outside the household | | (26) |
| $\beta_h$ | Probability of transmission per contact with an infectious individual within the household | See text | Computed within the model |
| $\beta_{h'}$ | Probability of transmission per contact with an infectious individual outside the household | See text | Computed within the model |
| $z_t$ | Proportion of people eligible for testing who are tested | As estimated in mass-testing (0.85, 0.78, 0.78) | Slovakia MOH (2020) |
| $c_p$ | Compliance with quarantine for those who test positive | 1.0 | Assumption |
| $c_h$ | Compliance with quarantine for household members of those who test positive | Variable: 0.0, 0.5, 1.0 | Scenario testing |
| $R_E^*$ | Target $R_E$ after implementation of extended contact reducing measures | Variable: 0.6, 1.0 | Scenario testing, based on the range of R estimates in November lockdowns in other EU countries (24) |
| $P_E$ | Sensitivity of SARS-CoV-2 test among individuals in latent class | 0 | SD Biosensor test does not detect infections when not infectious (13, 38) |
| $P_P$ | Sensitivity of SARS-CoV-2 test among individuals in pre-symptomatic infectious class | 100% | SD Biosensor test does not detect infections when not infectious (13, 38) |
| $P_C$ | Sensitivity of SARS-CoV-2 test among individuals in symptomatic infectious class | 100% | SD Biosensor test does not detect infections when not infectious (13, 38) |
| $P_S$ | Sensitivity of SARS-CoV-2 test among individuals in asymptomatic infectious class | 100% | SD Biosensor test does not detect infections when not infectious (13, 38) |

##### Simulations

We ran a total of 21 scenarios and 500 stochastic model iterations within each scenario:

Simulations We ran a total of 21 scenarios and 500 stochastic model iterations within each scenario:
| Scenario | Extended contact reducing measures effectiveness ($R_E^*$) | Number of test rounds | Compliance household members ($c_h$) |
| --- | --- | --- | --- |
| 1 | N/A | 0 | N/A |
| 2 | N/A | 1 | 100% |
| 3 | N/A | 2 | 100% |
| 4 | N/A | 3 | 100% |
| 5 | N/A | 1 | 50% |
| 6 | N/A | 2 | 50% |
| 7 | N/A | 3 | 50% |
| 8 | N/A | 1 | 0% |
| 9 | N/A | 2 | 0% |
| 10 | N/A | 3 | 0% |
| 11 | 1 | 0 | N/A |
| 12 | 1 | 1 | 100% |
| 13 | 1 | 2 | 100% |
| 14 | 1 | 3 | 100% |
| 15 | 1 | 1 | 50% |
| 16 | 1 | 2 | 50% |
| 17 | 1 | 3 | 50% |
| 18 | 1 | 1 | 0% |
| 19 | 1 | 2 | 0% |
| 20 | 1 | 3 | 0% |
| 21 | 0.6 | 0 | N/A |

## Literature

1. Guidelines for the implementation of non-pharmaceutical interventions against COVID-19. Eur. Cent. Dis. Prev. Control (2020), (available at https://www.ecdc.europa.eu/en/publications-data/covid-19-guidelines-non-pharmaceutical-interventions).

2. N. G. Davies, A. J. Kucharski, R. M. Eggo, A. Gimma, W. J. Edmunds, T. Jombart, K. O’Reilly, A. Endo, J. Hellewell, E. S. Nightingale, B. J. Quilty, C. I. Jarvis, T. W. Russell, P. Klepac, N. I. Bosse, S. Funk, S. Abbott, G. F. Medley, H. Gibbs, C. A. B. Pearson, S. Flasche, M. Jit, S. Clifford, K. Prem, C. Diamond, J. Emery, A. K. Deol, S. R. Procter, K. van Zandvoort, Y. F. Sun, J. D. Munday, A. Rosello, M. Auzenbergs, G. Knight, R. M. G. J. Houben, Y. Liu, Effects of non-pharmaceutical interventions on COVID-19 cases, deaths, and demand for hospital services in the UK: a modelling study. Lancet Public Health. 0 (2020), doi:10.1016/S2468-2667(20)30133-X.

3. S. Flaxman, S. Mishra, A. Gandy, H. J. T. Unwin, T. A. Mellan, H. Coupland, C. Whittaker, H. Zhu, T. Berah, J. W. Eaton, M. Monod, A. C. Ghani, C. A. Donnelly, S. Riley, M. A. C. Vollmer, N. M. Ferguson, L. C. Okell, S. Bhatt, Estimating the effects of non-pharmaceutical interventions on COVID-19 in Europe. Nature. 584, 257–261 (2020).

4. Everyone Included: Social Impact of COVID-19 | DISD, (available at https://www.un.org/development/desa/dspd/everyone-included-covid-19.html).

5. Socio-economic impact of COVID-19. UNDP, (available at https://www.undp.org/content/undp/en/home/coronavirus/socio-economic-impact-of-covid-19.html).

6. A. J. Kucharski, P. Klepac, A. J. K. Conlan, S. M. Kissler, M. L. Tang, H. Fry, J. R. Gog, W. J. Edmunds, CMMID COVID-19 working group, Effectiveness of isolation, testing, contact tracing, and physical distancing on reducing transmission of SARS-CoV-2 in different settings: a mathematical modelling study. Lancet Infect. Dis. 20, 1151–1160 (2020).

7. M. E. Kretzschmar, G. Rozhnova, M. C. J. Bootsma, M. van Boven, J. H. H. M. van de Wijgert, M. J. M. Bonten, Impact of delays on effectiveness of contact tracing strategies for COVID-19: a modelling study. Lancet Public Health. 5, e452–e459 (2020).

8. R. Wölfel, V. M. Corman, W. Guggemos, M. Seilmaier, S. Zange, M. A. Müller, D. Niemeyer, T. C. Jones, P. Vollmar, C. Rothe, M. Hoelscher, T. Bleicker, S. Brünink, J. Schneider, R. Ehmann, K. Zwirglmaier, C. Drosten, C. Wendtner, Virological assessment of hospitalized patients with COVID-2019. Nature. 581, 465–469 (2020).

9. M. J. Mina, R. Parker, D. B. Larremore, Rethinking Covid-19 Test Sensitivity — A Strategy for Containment. N. Engl. J. Med. (2020), doi:10.1056/NEJMp2025631.

10. D. B. Larremore, B. Wilder, E. Lester, S. Shehata, J. M. Burke, J. A. Hay, M. Tambe, M. J. Mina, R. Parker, Test sensitivity is secondary to frequency and turnaround time for COVID-19 screening. Sci. Adv. 7, eabd5393 (2021).

11. Oxford University, “An observational study of SARS-CoV-2 infectivity by viral load and demographic factors and the utility lateral flow devices to prevent transmission,” (available at https://www.ox.ac.uk/news/2021-01-21-lateral-flow-devices-detect-most-infectious-covid-19-cases-and-could-allow-safer).

12. Som zodpovedny. Som Zodp., (available at https://www.somzodpovedny.sk/).

13. Z. Iglὁi, J. Velzing, J. van Beek, D. van de Vijver, G. Aron, R. Ensing, K. Benschop, W. Han, T. Boelsums, M. Koopmans, C. Geurtsvankessel, R. Molenkamp, medRxiv, in press, doi:10.1101/2020.11.18.20234104.

14. K. Bodová, R. Kollár, medRxiv, in press, doi:10.1101/2020.12.23.20248808.

15. S. Riley, O. Eales, C. E. Walters, H. Wang, K. E. C. Ainslie, C. Atchison, C. Fronterre, P. J. Diggle, D. Ashby, C. A. Donnelly, G. Cooke, W. Barclay, H. Ward, A. Darzi, P. Elliott, medRxiv, in press, doi:10.1101/2020.11.30.20239806.

16. Panbio COVID-19 Ag Rapid Test Device, (available at https://www.globalpointofcare.abbott/en/product-details/panbio-covid-19-ag-antigen-test.html).

17. Sofia SARS Antigen FIA | Quidel, (available at https://www.quidel.com/immunoassays/rapid-sars-tests/sofia-sars-antigen-fia).

18. E. Mahase, Covid-19: Innova lateral flow test is not fit for “test and release” strategy, say experts. BMJ. 371 (2020), doi:10.1136/bmj.m4469.

19. How Slovakia tested 3.6 million people for COVID-19 in a single weekend, (available at https://www.bi.team/blogs/how-slovakia-tested-3-6-million-people-for-covid-19-in-a-single-weekend/).

20. Products - STANDARD Q COVID-19 Ag, (available at http://sdbiosensor.com/xe/product/7672).

21. A. Berger, M. T. N. Nsoga, F. J. Perez-Rodriguez, Y. A. Aad, P. Sattonnet-Roche, A. Gayet-Ageron, C. Jaksic, G. Torriani, E. Boehm, I. Kronig, J. A. Sacks, M. de Vos, F. J. Bausch, F. Chappuis, L. Kaiser, M. Schibler, I. Eckerle, for the G. C. for E. V. Diseases, medRxiv, in press, doi:10.1101/2020.11.20.20235341.

22. J. L. Prince-Guerra, O. Almendares, L. D. Nolen, J. K. L. Gunn, A. P. Dale, S. A. Buono, M. Deutsch-Feldman, S. Suppiah, L. Hao, Y. Zeng, V. A. Stevens, K. Knipe, J. Pompey, C. Atherstone, D. P. Bui, T. Powell, A. Tamin, J. L. Harcourt, P. L. Shewmaker, M. Medrzycki, P. Wong, S. Jain, A. Tejada-Strop, S. Rogers, B. Emery, H. Wang, M. Petway, C. Bohannon, J. M. Folster, A. MacNeil, R. Salerno, W. Kuhnert-Tallman, J. E. Tate, N. J. Thornburg, H. L. Kirking, K. Sheiban, J. Kudrna, T. Cullen, K. K. Komatsu, J. M. Villanueva, D. A. Rose, J. C. Neatherlin, M. Anderson, P. A. Rota, M. A. Honein, W. A. Bower, Evaluation of Abbott BinaxNOW Rapid Antigen Test for SARS-CoV-2 Infection at Two Community-Based Testing Sites — Pima County, Arizona, November 3–17, 2020. MMWR Morb. Mortal. Wkly. Rep. 70 (2021), doi:10.15585/mmwr.mm7003e3.

23. Koronavírus a Slovensko. Koronavírus Slov., (available at https://korona.gov.sk/).

24. S. Abbott, J. Hellewell, R. N. Thompson, K. Sherratt, H. P. Gibbs, N. I. Bosse, J. D. Munday, S. Meakin, E. L. Doughty, J. Y. Chun, Y.-W. D. Chan, F. Finger, P. Campbell, A. Endo, C. A. B. Pearson, A. Gimma, T. Russell, CMMID COVID modelling group, S. Flasche, A. J. Kucharski, R. M. Eggo, S. Funk, Estimating the time-varying reproduction number of SARS-CoV-2 using national and subnational case counts. Wellcome Open Res. 5, 112 (2020).

25. Statistics | Eurostat, (available at https://ec.europa.eu/eurostat/databrowser/view/cens_01rhsize/default/table?lang=en).

26. K. Prem, K. van Zandvoort, P. Klepac, R. M. Eggo, N. G. Davies C. for the M. M. of I. D. C.-19 W. Group, A. R. Cook, M. Jit, medRxiv, in press, doi:10.1101/2020.07.22.20159772.

27. R Core Team, R: A Language and Environment for Statistical Computing (R Foundation for Statistical Computing, Vienna, Austria, 2019; https://www.R-project.org/).

28. United Nations, “World Population prospects,” (available at https://www.google.com/url?q=‖whttps://population.un.org/wpp/Download/Files/1_Indicators%20(Standard)/EXCEL_FILES/1_Population/WPP2019_POP_F01_1_TOTAL_POPULATION_BOTH_SEXES.xlsx&sa=D&source=editors&ust=1614209701533000&usg=AOvVaw0e_BV1Z-ofSu_U0Q5NeZBf).

29. European Commission, Eurostat Sata Browser (https://ec.europa.eu/eurostat/databrowser/view/CENS_01RHSIZEcustom_214489/default/table?lang=en).

30. S. A. Lauer, K. H. Grantz, Q. Bi, F. K. Jones, Q. Zheng, H. R. Meredith, A. S. Azman, N. G. Reich, J. Lessler, The Incubation Period of Coronavirus Disease 2019 (COVID-19) From Publicly Reported Confirmed Cases: Estimation and Application. Ann. Intern. Med. (2020), doi:10.7326/M20-0504.

31. L. Ferretti, A. Ledda, C. Wymant, L. Zhao, V. Ledda, L. Abeler-Dörner, M. Kendall, A. Nurtay, H.-Y. Cheng, T.-C. Ng, H.-H. Lin, R. Hinch, J. Masel, A. M. Kilpatrick, C. Fraser, medRxiv, in press, doi:10.1101/2020.09.04.20188516.

32. Q. Bi, Y. Wu, S. Mei, C. Ye, X. Zou, Z. Zhang, X. Liu, L. Wei, S. A. Truelove, T. Zhang, W. Gao, C. Cheng, X. Tang, X. Wu, Y. Wu, B. Sun, S. Huang, Y. Sun, J. Zhang, T. Ma, J. Lessler, T. Feng, Epidemiology and transmission of COVID-19 in 391 cases and 1286 of their close contacts in Shenzhen, China: a retrospective cohort study. Lancet Infect. Dis., S1473309920302875 (2020).

33. H. Nishiura, N. M. Linton, A. R. Akhmetzhanov, Serial interval of novel coronavirus (COVID-19) infections. Int. J. Infect. Dis. 93, 284–286 (2020).

34. Q. Li, X. Guan, P. Wu, X. Wang, L. Zhou, Y. Tong, R. Ren, K. S. M. Leung, E. H. Y. Lau, J. Y. Wong, X. Xing, N. Xiang, Y. Wu, C. Li, Q. Chen, D. Li, T. Liu, J. Zhao, M. Liu, W. Tu, C. Chen, L. Jin, R. Yang, Q. Wang, S. Zhou, R. Wang, H. Liu, Y. Luo, Y. Liu, G. Shao, H. Li, Z. Tao, Y. Yang, Z. Deng, B. Liu, Z. Ma, Y. Zhang, G. Shi, T. T. Y. Lam, J. T. Wu, G. F. Gao, B. J. Cowling, B. Yang, G. M. Leung, Z. Feng, Early Transmission Dynamics in Wuhan, China, of Novel Coronavirus–Infected Pneumonia. N. Engl. J. Med. 382, 1199–1207 (2020).

35. N. G. Davies, P. Klepac, Y. Liu, K. Prem, M. Jit, R. M. Eggo, Age-dependent effects in the transmission and control of COVID-19 epidemics. Nat. Med. 26, 1205–1211 (2020).

36. Y. Liu, Centre for Mathematical Modelling of Infectious Diseases nCoV Working Group, S. Funk, S. Flasche, The contribution of pre-symptomatic infection to the transmission dynamics of COVID-2019. Wellcome Open Res. 5, 58 (2020).

37. X. He, E. H. Y. Lau, P. Wu, X. Deng, J. Wang, X. Hao, Y. C. Lau, J. Y. Wong, Y. Guan, X. Tan, X. Mo, Y. Chen, B. Liao, W. Chen, F. Hu, Q. Zhang, M. Zhong, Y. Wu, L. Zhao, F. Zhang, B. J. Cowling, F. Li, G. M. Leung, Temporal dynamics in viral shedding and transmissibility of COVID-19. Nat. Med. 26, 672–675 (2020).

38. L. J. Krüger, M. Gaeddert, L. Köppel, L. E. Brümmer, C. Gottschalk, I. B. Miranda, P. Schnitzler, H. G. Kräusslich, A. K. Lindner, O. Nikolai, F. P. Mockenhaupt, J. Seybold, V. M. Corman, C. Drosten, N. R. Pollock, A. I. Cubas-Atienzar, K. Kontogianni, A. Collins, A. H. Wright, B. Knorr, A. Welker, M. de Vos, J. A. Sacks, E. R. Adams, C. M. Denkinger F. the S. Team, medRxiv, in press, doi:10.1101/2020.10.01.20203836.

